# A role for super-spreaders in carrying malaria parasites across the months-long dry season

**DOI:** 10.1101/2022.04.28.22274398

**Authors:** Eva Stadler, Deborah Cromer, Samson Ogunlade, Aissata Ongoiba, Safiatou Doumbo, Kassoum Kayentao, Boubacar Traore, Peter D Crompton, Silvia Portugal, Miles P Davenport, David S Khoury

## Abstract

In malaria endemic regions, transmission of *Plasmodium falciparum* parasites is often seasonal with very low transmission during the dry season and high transmission in the wet season. Parasites survive the dry season within some individuals who experience prolonged carriage of parasites and are thought to ‘seed’ infection in the next transmission season. We use a combination of mathematical simulations and data analysis to characterise dry season carriers and their role in the subsequent transmission season. Simulating the life-history of individuals experiencing repeated exposure to infection predicts that dry season carriage is more likely in the oldest, most exposed and most immune individuals. This hypothesis is supported by data from a longitudinal study in Mali that shows that carriers are significantly older, experience a higher biting rate at the beginning of the transmission season and develop clinical malaria later than non-carriers. Further, since the most exposed individuals in a community are most likely to be dry season carriers, we show that this is predicted to enable a more than 2-fold faster spread of parasites into the mosquito population at the start of the subsequent wet season.

## Introduction

Malaria caused an estimated 627,000 deaths worldwide in 2020 (World Health Organization, 2021). It is caused by *Plasmodium* parasites, the most prevalent being *Plasmodium falciparum* and *Plasmodium vivax*. Seasonal transmission of *Plasmodium* parasites, that is, low transmission during the dry season and higher transmission during the wet season, is common in malaria endemic regions (Druilhe & Pérignon, 1997; Jawara et al., 2008). During the dry season, the transmission rate is very low, in some areas it is near zero (Lehmann et al., 2010; Portugal et al., 2017). This raises the question of how *P. falciparum* parasites survive the dry season and re-seed infection in the wet season? In malaria endemic regions, adults are often asymptomatic, but harbor low parasite loads (Babiker, Abdel-Muhsin, Ranford-Cartwright, Satti, & Walliker, 1998; Baum et al., 2015; Druilhe & Pérignon, 1997; Imwong et al., 2016; Roper et al., 1996). These sub-patent malaria infections have also been found during the dry season (Andrade et al., 2020; Babiker et al., 1998; Imwong et al., 2016; Roper et al., 1996) and have led to the conclusion that humans provide a reservoir for parasites during the dry season (Babiker et al., 1998; Jawara et al., 2008; Struik & Riley, 2004). Recently, Andrade et al. have shown that parasites survive at low levels in certain individuals and carriers have higher humoral immunity and protection against clinical malaria than non-carriers (Andrade et al., 2020) as was also shown by Portugal et al. (Portugal et al., 2017). At the same study site only 12 to 30% of individuals carried *P. falciparum* parasites at the end of the dry season in different years (Andrade et al., 2020; Portugal et al., 2017). This small subset of individuals will likely contribute to the infection of the vector population at the commencement of the wet season and thereby re-seed the next transmission season. This is supported by studies showing that the genetic diversity of parasites is preserved in the dry season (Andrade et al., 2020; Babiker et al., 1998; Portugal et al., 2017). However, it remains unclear whether this minority of individuals who carry infection is a random subset of the individuals exposed to infection in the previous season or if they have some, as yet unidentified, factor contributing to their propensity to carry parasites. Thus, we study what characterises these dry season carriers.

Age appears to be an important correlate of carriage (Selvaraj, Wenger, & Gerardin, 2018). In high transmission regions, age is both a determinant of the parasite density distribution (Imwong et al., 2016) and cumulative past exposure (Felger et al., 2012). Older children and adults have been shown to have longer lasting infections as well as higher immunity (Doolan, Dobaño, & Baird, 2009; Douglas et al., 2011; Recker & Gupta, 2006; Rodriguez-Barraquer et al., 2018; Smith & Vounatsou, 2003). Previous modelling work has hypothesized a mechanism by which increased immunity as a result of age and exposure may drive longer durations of infection (Pinkevych et al., 2012; Recker & Gupta, 2006; Smith & Vounatsou, 2003). This may occur due to antibody-mediated cross-reactive immunity that controls diverse *P. falciparum* strains, but it may also be due to a more general physiological response such as hypersplenism (Pinkevych et al., 2012) or changes in the activation of endothelial cells and reduced sequestration, which lead to overall slower growing parasitemia (Andrade et al., 2020; Azasi et al., 2018; David, Hommel, Miller, Udeinya, & Oligino, 1983). Regardless of the mechanism, modelling has suggested that if exposure to infection induces immunity, longer-lived infections will occur as an individual’s exposure increases. In addition to age, individuals may also vary in their degree of exposure (Cooper et al., 2019; Selvaraj et al., 2018). For this reason, some may acquire a level of immunity sufficient for long-lived infections earlier in life than others. Therefore, here we explore the role of exposure to infection in determining parasite carriage across the dry season.

We first assess the relationship between exposure, age, and parasite carriage using a mathematical model of repeated infections within an individual and explore the stochastic variation in an individual’s propensity to carry infection. By comparing with cohort data from Mali (Portugal et al., 2017; Tran et al., 2013), we then explore whether this stochasticity is sufficient to explain why some individuals carry parasites over the dry seasons and others do not. We observe in this cohort data that parasite carriage is not purely stochastic and is in fact more likely in individuals who are most at risk of infection. The highly exposed individuals who carry parasites over the dry season, if they are bitten more frequently, are likely to re-seed infection at the end of the wet season and predicted to cause a 2.6-fold faster infection of the mosquito population than if carriage of infection over the dry-season is from a random subset of individuals. This suggests that dry-season carriers may play a role as “super-spreaders” in seasonal transmission settings.

## Results

### Simulation demonstrates a random subset of individuals can carry infection over the dry season

First, we used a simulation model based on a model by Pinkevych et al. (Pinkevych et al., 2012) to explore how exposure and age influence the probability of an individual carrying infection over the dry season (***Figure 1*** and Materials and Methods). This model simulates an individual’s exposure to repeated infections. Infectious bites are stochastic and induce both strain specific immune responses and cross-reactive immune responses. Each infectious bite is assumed to be a different strain, and cross-reactive immunity decays with a 5 year half-life. We simulated individuals’ first 20 years of infection histories under different Forces Of Infection (FOIs) (***Figure 2, Supplementary Fig. S1***, and ***Supplementary Fig. S2***). After an infectious mosquito bite with a particular strain, the parasite concentration increases which induces an increase in strain specific immunity and a slower increase in cross-reactive immunity (***Figure 2***A, ***B***). High parasite loads in early childhood infection in the model induces large numbers of antibodies in the model. Thus, the less well controlled an infection is, the higher the peak of strain specific immune response (***Figure 2***). This result follows from the model assumption that antibody production is correlated with pathogen load. It follows that the first infections are predominantly controlled by strain specific immunity, but as the individual experiences repeated infections, cross-reactive immunity has a much greater contribution to the control of infections (***Figure 2***B). Higher levels of cross-reactive immunity lead to a decrease in the maximal parasite concentration, and in turn lower peak strain specific immune responses. A property that emerges from the model is that as cross-reactive immunity increases, the lower peak in parasite concentration and strain specific immunity lead to a longer duration of each infection (***Figure 2***C, ***D***). We performed each simulation 10,000 times for each FOI tested. These results revealed that the age of an individual first carrying parasites over the dry season decreases nonlinearly with increasing FOI (***Supplementary Fig. S3*** and ***Supplementary Fig. S4***). Moreover, the probability of an individual carrying parasites is higher with higher FOIs (***Figure 3***). These simulations reveal that for a given exposure and age, there is a certain chance an individual will carry parasites over the dry season, and a chance they will not. We next explored whether any factor predicted whether an individual would carry parasites over a dry season.

**Figure 1.**
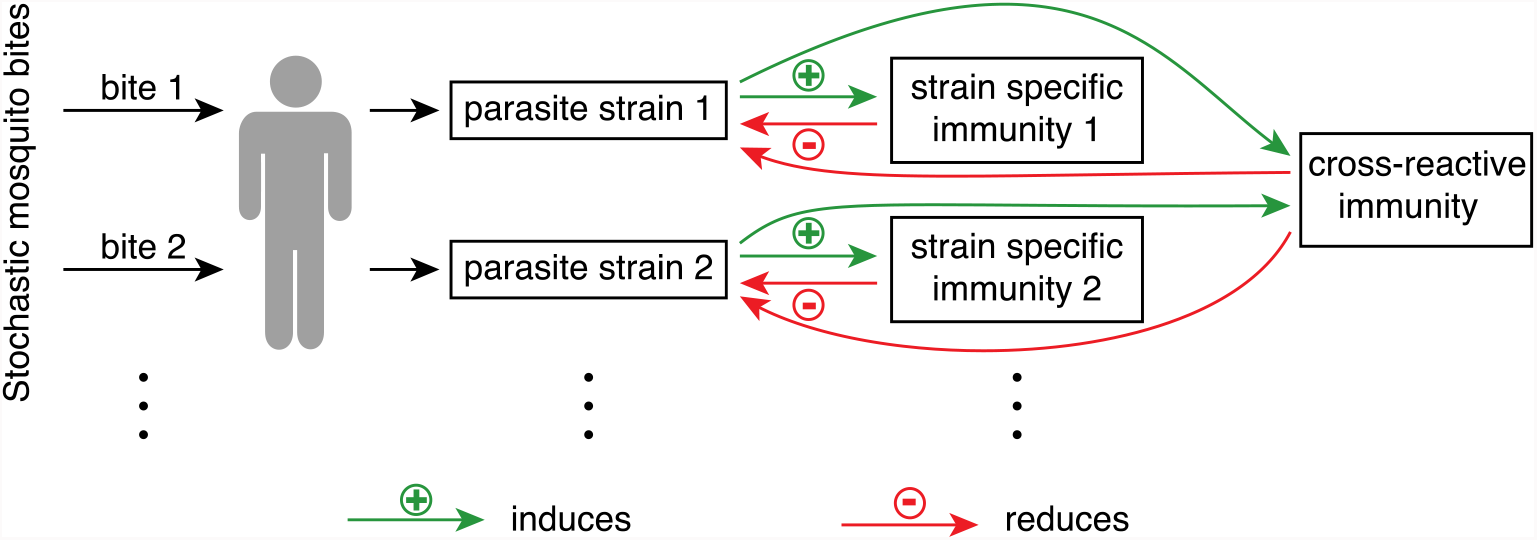
Schematic of the mathematical model used to simulate the malaria life history of individuals. Individuals receive stochastic and seasonal infectious mosquito bites. Each bite inoculates a new parasite strain. The growth of a parasite strain induces strain specific immunity to that strain and cross-reactive immunity to all strains. Immunity reduces the growth rate of either a specific strain (strain specific immunity) or all strains (cross-reactive immunity). If the parasite concentration is low, cross-reactive immunity decreases whereas strain specific immunity only decreases when the corresponding parasite strain is cleared. More information on the model can be found in the Materials and Methods and the supplementary methods. Representative simulations of individuals with different exposure to infectious mosquito bites are shown in ***Figure 2, Supplementary Fig. S1*** and ***Supplementary Fig. S2***.

**Figure 2.**
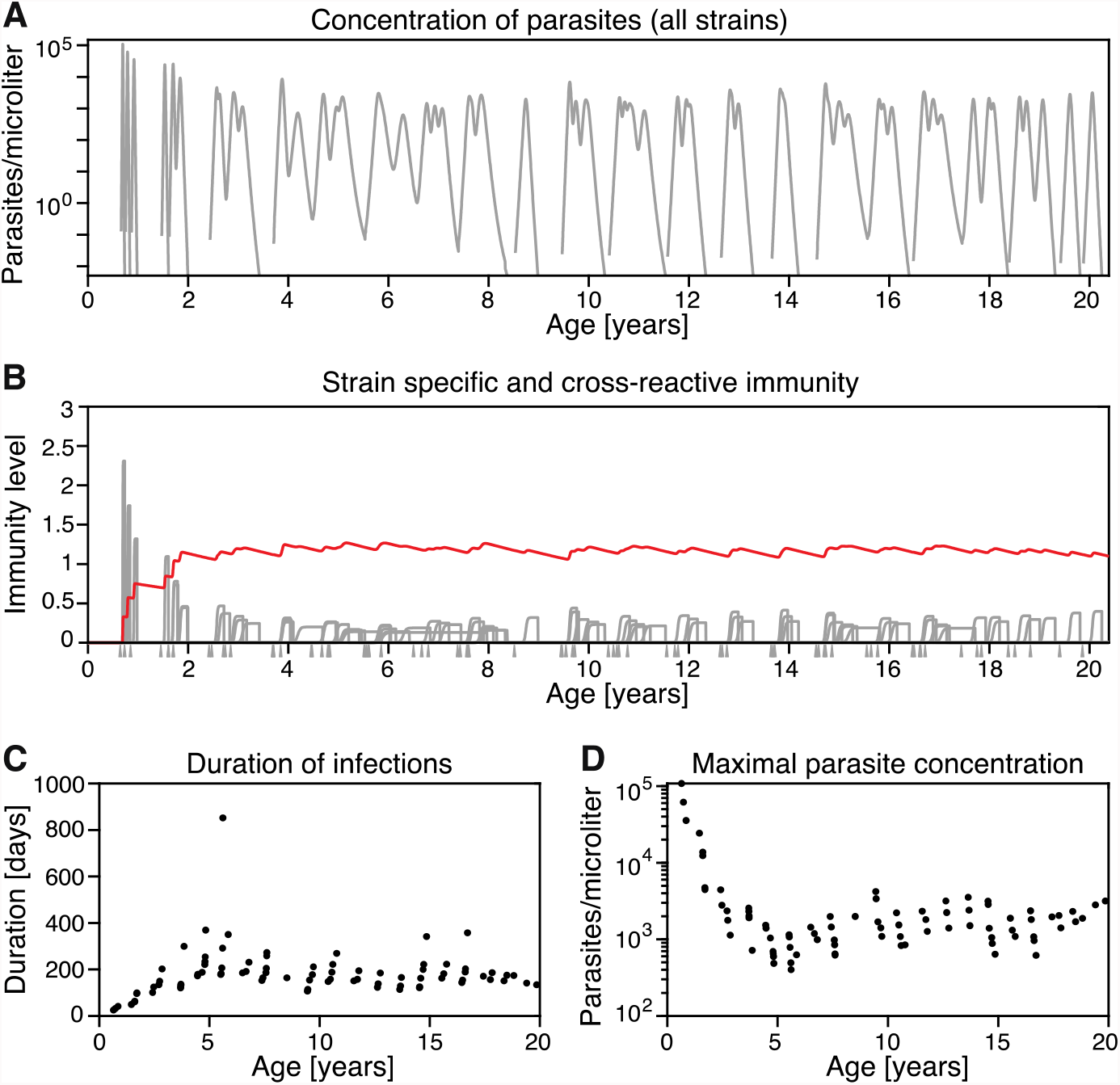
Simulation of one individual from birth to age 20. We simulated an individual with a mean biting rate of 0.022 bites per day (all other parameters as in ***Supplementary Table S1***). (**A**) Overall parasite concentration, i.e., the concentration of all parasite strains together, with age. (**B**) Strain specific immunity (grey lines) and cross-reactive immunity (red line) vs age. Cross-reactive immunity may be antibody-mediated cross-reactive immunity or a more cross-reactive physiological response. The grey triangles indicate infectious mosquito bites. Note that strain specific immunity is only plotted for the time interval the respective strain is present in the blood. (**C**) Duration of infections with age. The duration of an infection is the time from the beginning of the blood-stage to clearance of the parasite strain. (**D**) Peak parasite concentration of each infection that occurs in the first 20 years of an individual’s life from the simulation, i.e., the maximal parasite concentration for each infection. As the individual generates more cross-reactive immunity with age and exposure, the peak parasite concentrations experienced by that individual decline. For simulations of individuals with a different exposure, see ***Supplementary Fig. S1*** and ***Supplementary Fig. S2***.

**Figure 3.**
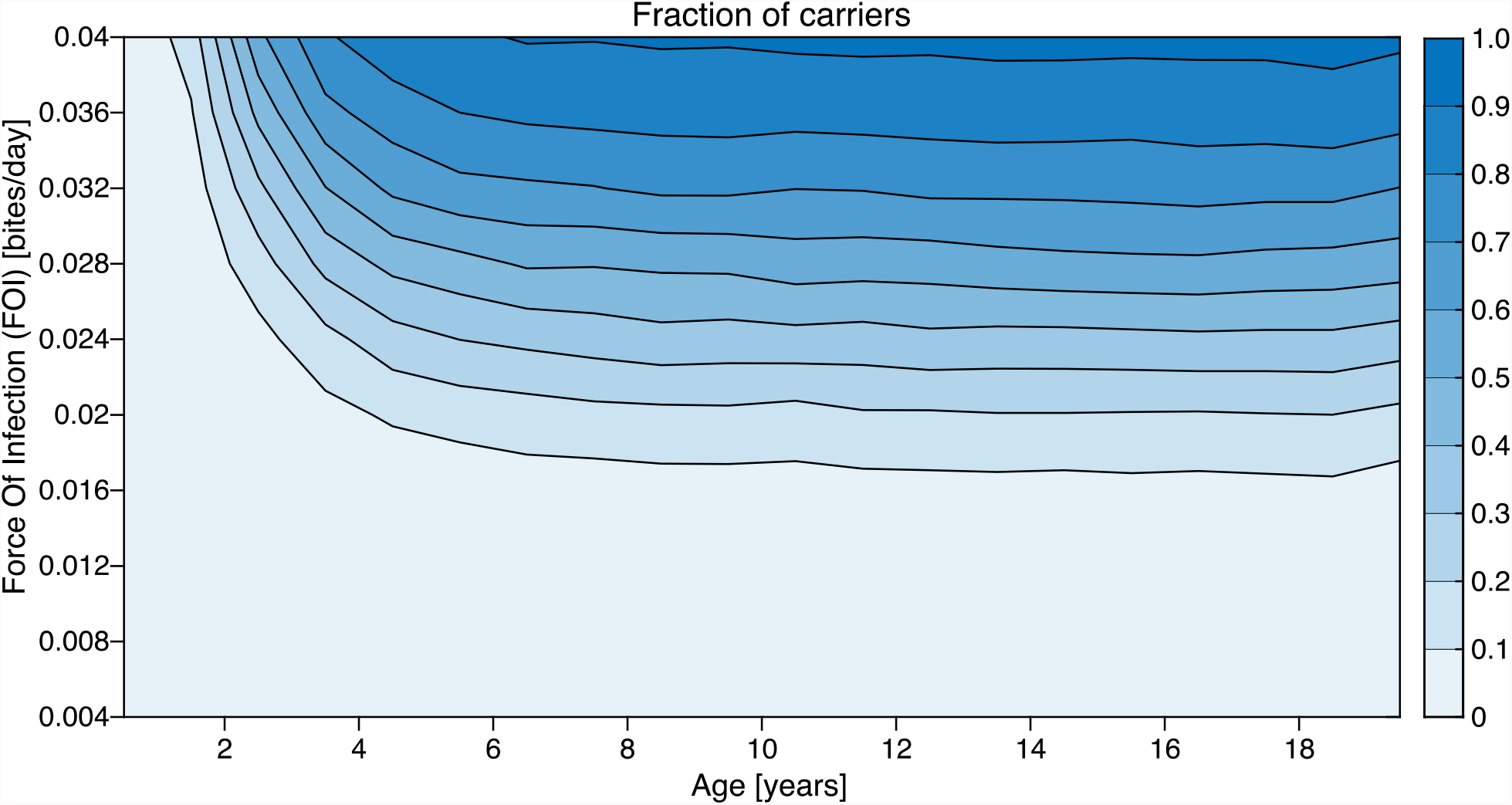
Fraction of simulated individuals who carry parasites over the dry season by FOI and age. For each of ten different FOIs between 0.004 and 0.04 bites per day during the wet season, 10,000 individuals were simulated from birth to age 20. The different shaded areas indicate which fraction of the simulated individuals has carried parasites through the dry season at the respective age. We define parasite carriage as PCR detectable parasite concentrations in an individual on the last day of the dry season. Individuals with a higher exposure carry parasites at a younger age, while some individuals who are less exposed never carry parasites over the dry season before they reach age 20 (see also ***Supplementary Fig. S3*** and ***Supplementary Fig. S4***).

For a given FOI, we found that individuals who carried parasites over a dry season had experienced significantly more infectious bites and a more recent infectious bite in the previous wet season than individuals who did not carry parasites (***Figure 4, Supplementary Fig. S5*** and ***Supplementary Fig. S6***). Together these modelling results predict that a history of high exposure to infection may increase the risk of carriage over the dry season and that parasite carriage may be purely random. That is, the same individual may or may not carry parasites over a dry season depending on their recent infection history.

**Figure 4.**
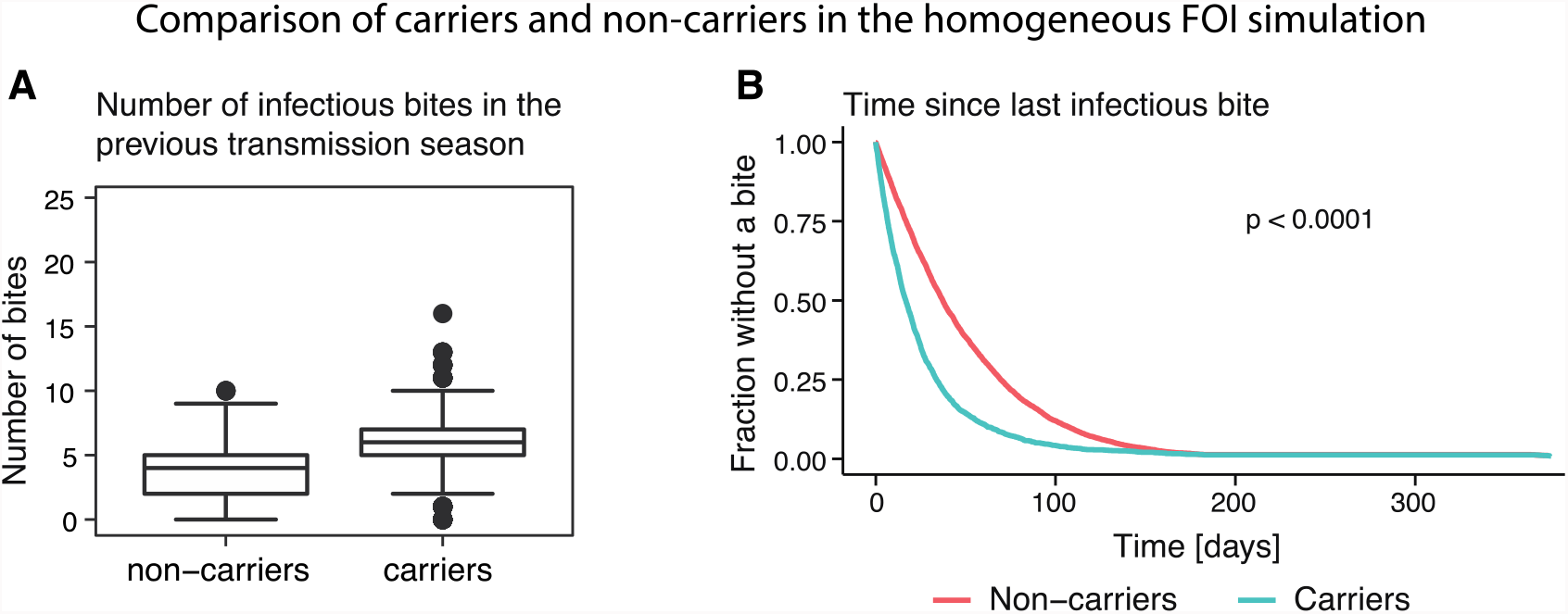
Simulation of individuals with a FOI of 0.024 bites per day. For this simulation, 10,000 individuals with the same exposure were simulated to an age uniformly drawn between 0 and 20 years. Individuals were classified as carriers if they had a parasite concentration above the limit of detection for a Rapid Diagnostic Test (RDT) at the end of the dry season. (**A**) The number of infectious bites that individuals received in the transmission season before they were classified as carriers or non-carriers is significantly higher for carriers than for non-carriers (Wilcoxon rank-sum test with p-value < 0.0001). (**B**) The time from the last infectious bite to the end of the previous transmission season is significantly shorter for carriers than for non-carriers (log-rank test with p-value < 0.0001). For a comparison of the number of infectious bites in the previous season and the time since the last infectious bite between carriers and non-carriers for other FOIs see ***Supplementary Fig. S5*** and ***Supplementary Fig. S6***.

### Time to next infection distinguishes stochastic and exposure-mediated dry season carriage

The above modelling demonstrated that purely stochastic differences are a possible reason why a fraction of individuals with similar immunity and history of exposure may carry parasites over the dry season while others do not, and there may be no distinctive characteristics of the carrier population. However, since the probability of dry season parasite carriage increases with the FOI (***Figure 3***), it is also possible that dry season carriers are not just a random subset of individuals in a community but that they are the most frequently infected. We now simulate two possible scenarios: 1) a community of individuals exposed to a homogenous FOI, and 2) a community of individuals exposed to a heterogenous FOI. In these two scenarios, we identify and characterise the subset of individuals who carried parasites over the dry season. These two scenarios show some features that are consistent. In both the homogeneous and the heterogeneous exposure settings, carriers are significantly older and more immune than non-carriers (***Figure 5***C-**F, *Supplementary Table S2, Supplementary Fig. S8*** and ***Supplementary Fig. S11***). The difference between the homogeneous and the heterogeneous simulation is that in the latter, carriers are more exposed than non-carriers (***Figure 5***H, ***Supplementary Fig. S8*** and ***Supplementary Fig. S11***) while in the homogeneous case there is no difference in the exposure of carriers and non-carriers (***Supplementary Table S2***). Interestingly, despite the systematically higher risk of infection between carriers and non-carriers in the heterogeneous population model, we find that there is also stochasticity in the heterogeneous case – while older and more exposed individuals are more likely to be carriers, some younger or less exposed individuals may also be carriers (***Supplementary Fig. S9***) and it is not necessarily the same individuals that carry parasites over different dry seasons (***Supplementary Fig. S10***). Together, this modelling predicts that in a heterogenous population a subset of individuals with higher risk of infection are more likely to carry parasites over the dry season.

**Figure 5.**
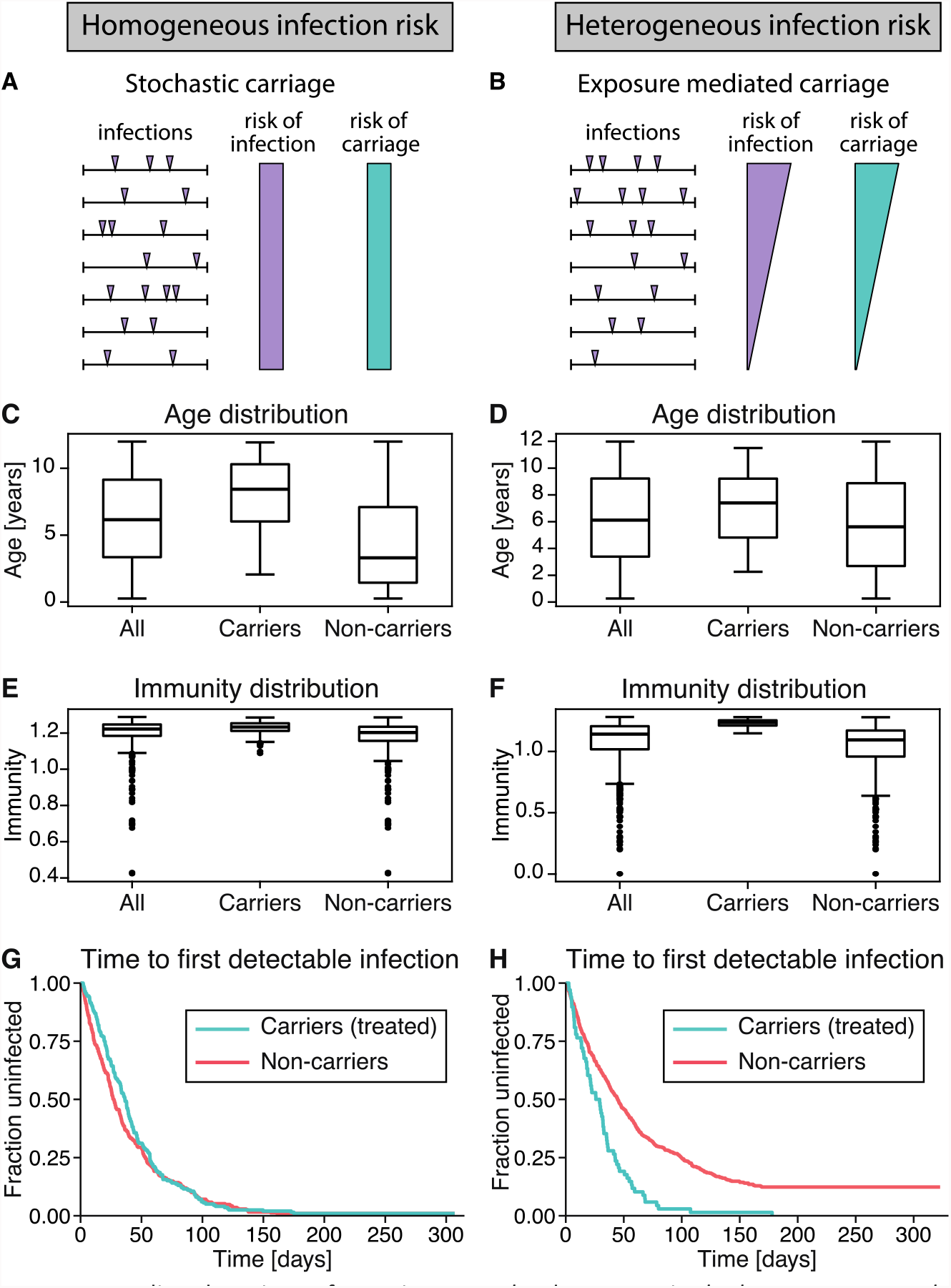
Stochastic or exposure mediated carriage of parasites over the dry season in the homogeneous or heterogeneous exposure settings, respectively. (**A**) If parasite carriage over the dry season is stochastic in a homogeneous population, then all individuals have a similar risk of infection and risk of carriage. (**B**) If parasite carriage is exposure mediated, then the individuals with a higher risk of infection also have a higher risk of parasite carriage over the dry season. In this scenario, there is still stochasticity in carriage but the risk of carriage that correlates with the risk of infection. (**C**-**H**) We simulated 1,000 individuals aged between 3 months and 12 years with either homogeneous or heterogeneous infection risk during the wet season. Individuals were classified as carriers or non-carriers depending on whether they have a parasite concentration above the limit of detection for a Rapid Diagnostic Test (RDT) at the end of the dry season. The simulation included treatment of carriers at the end of the dry season. Parameter values for this simulation can be found in ***Supplementary Table S1***. The biting rate for the homogeneous infection risk simulation is 0.024 bites per day (for simulations of a homogeneous population with other biting rates see ***Supplementary Table S2*** and ***Supplementary Fig. S7***). (**C, D**) In both the homogeneous and heterogeneous simulations, carriers were significantly older than non-carriers (Wilcoxon rank-sum test with p-values <0.0001 and 0.0005, respectively). (**E, F**) Carriers have a significantly higher cross-reactive immunity than non-carriers in both the homogeneous and heterogeneous infection risk simulation (Wilcoxon rank-sum test with p-values < 0.0001 in both cases). (**G, H**) Time from the first follow-up to infection (by PCR, i.e., the overall parasite concentration exceeds the limit of detection for PCR). In the simulation with homogeneous risk of infection, there is no significant difference in the time to the next infection between carriers and non-carriers (log-rank test with p-value 0.2). In the heterogeneous infection risk simulation, carriers had a significantly higher infection risk compared to non-carriers (log-rank test with p-value < 0.0001).

### Heterogeneity in infection risk in Mali cohort indicates that high risk of infection leads to high risk of dry season carriage

In the above we showed that a systematic difference of higher exposure or random differences may explain why some individuals carry infections over the dry season and others do not. Here, we investigate this model prediction in data from a seasonal malaria setting (Portugal et al., 2017; Tran et al., 2013). In particular, we consider whether there is any evidence that dry season carries are systematically more highly exposed individuals as previously hypothesized (Andrade et al., 2020; Portugal et al., 2017). In this cohort study (previously published by Tran et al. (Tran et al., 2013) and Portugal et al. (Portugal et al., 2017)), children aged 3 months to 12 years were tested for *P. falciparum* infection at the end of the dry season (using RDT and PCR) and those who were infected (RDT positive) were treated (***Figure 6***). Children were then followed up for infection (by PCR) fortnightly to the end of the year. Individuals who were positive for infection with a positive RDT result at the end of the dry season are assumed to be dry season carriers, while individuals without parasites (PCR and RDT negative) are assumed to be non-carriers (see Materials and Methods for more details on the data). These studies previously reported that carriers are more immune to clinical malaria than non-carriers (Andrade et al., 2020; Portugal et al., 2017), antibody levels decline similarly in carriers and non-carriers (Andrade et al., 2020), older children are more likely to be carriers (Portugal et al., 2017), and risk of clinical malaria decreases with age despite no age-related differences in the infection risk (Tran et al., 2013). Under both modelling scenarios older individuals with higher cross-reactive immunity were more likely to carry parasites over the dry seasons. Consistent with this in the cohort study, it has previously been shown that carriers were significantly older than non-carriers (data reproduced, ***Supplementary Fig. S12***) (Andrade et al., 2020; Portugal et al., 2017), and carriers had a lower risk of clinical malaria than non-carriers (data reproduced here, ***Figure 7***B, p-value < 0.0001) (Crompton et al., 2008; Doumbo et al., 2014; Portugal et al., 2017). Thus, consistent with our model, these previous studies indicate that carriers have a greater immunity to clinical malaria than non-carriers (Crompton et al., 2008; Doumbo et al., 2014; Portugal et al., 2017). Finally, to determine whether carriers had a systematically higher FOI, we performed a novel analysis of the Mali data, and compared the time to infection (by PCR) for carriers and non-carriers. Using both a log-rank test and Cox Proportional Hazards model to compare the time to infection data for carrier and non-carriers, we find that carriers had a significantly higher risk of infection compared to non-carriers, even after adjusting for age (***Figure 7***A, log-rank test with p-value < 0.0001, see also ***Supplementary Fig. S13*** and ***Supplementary Fig. S16***, Cox model with p<0.0001 for both age and parasite carriage, see ***Supplementary Table S3*** and ***Supplementary Fig. S15***, also see ***Supplementary Table S4, Supplementary Fig. S17, Supplementary Table S5*** and ***Supplementary Fig. S18***). Thus, we find that those who carry parasites at the end of the dry season have a higher risk of infection at the beginning of the wet season compared with non-carriers. This indicates that dry season carriage is not a purely random process and that carriers have a systemically higher exposure to infection.

**Figure 6.**
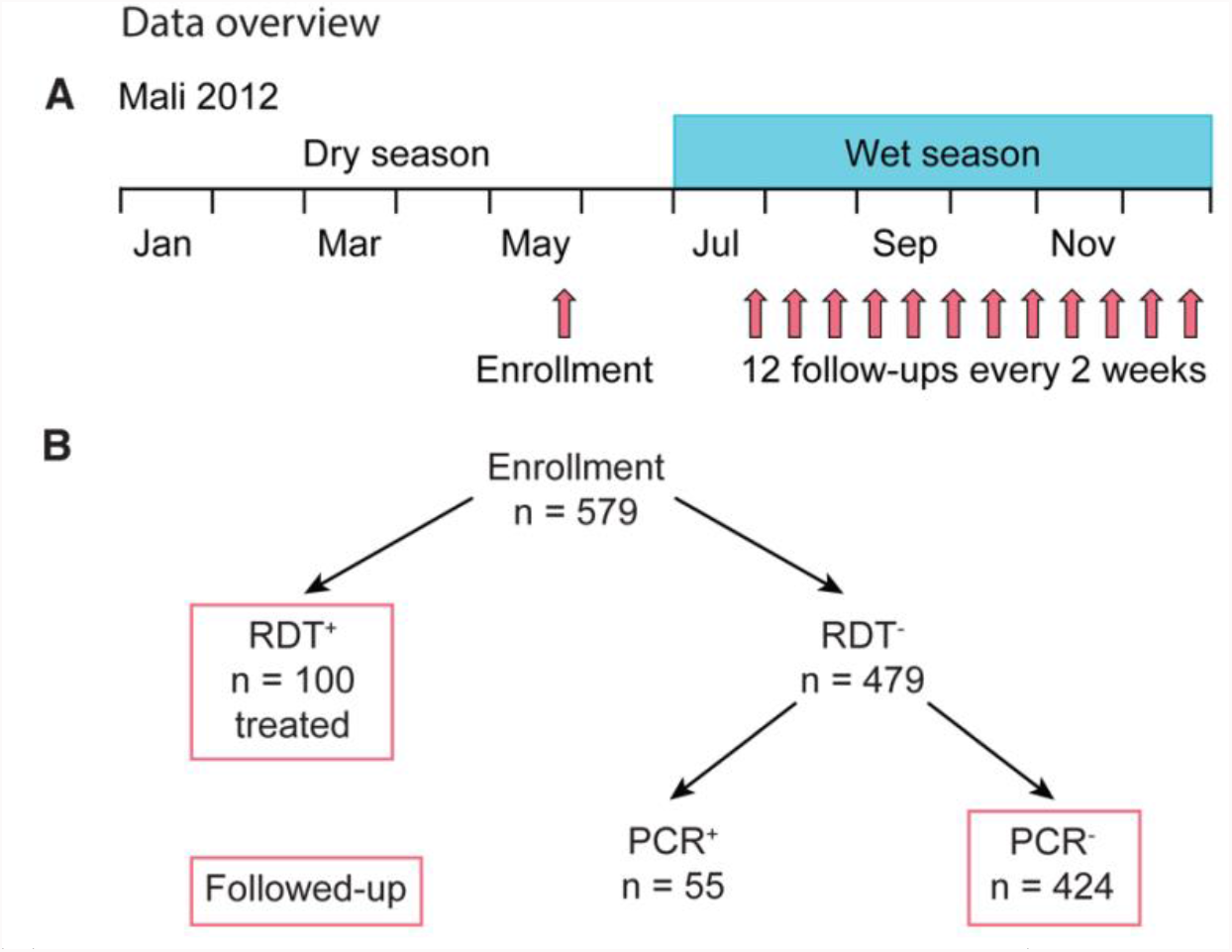
Data overview. (**A**) Children were enrolled in May 2012 at the end of the dry season (January to June). During the wet season, when malaria transmission takes place, enrolled children were visited every 14 days to the end of the year. Note that due to the political situation in Mali in 2012, there is a gap of two months between enrolment which took place in May 2012 and the first follow-up visit in July 2012. In order to take this gap in the data into account, we did the data analysis both with enrolment and the first follow-up visit as first visit date (see Supplementary information) and found the same results. (**B**) At enrolment, children were tested with RDTs. Those with positive test results were treated and those with negative results were additionally tested with PCR. Only RDT positive and RDT and PCR negative children were followed-up regularly.

**Figure 7.**
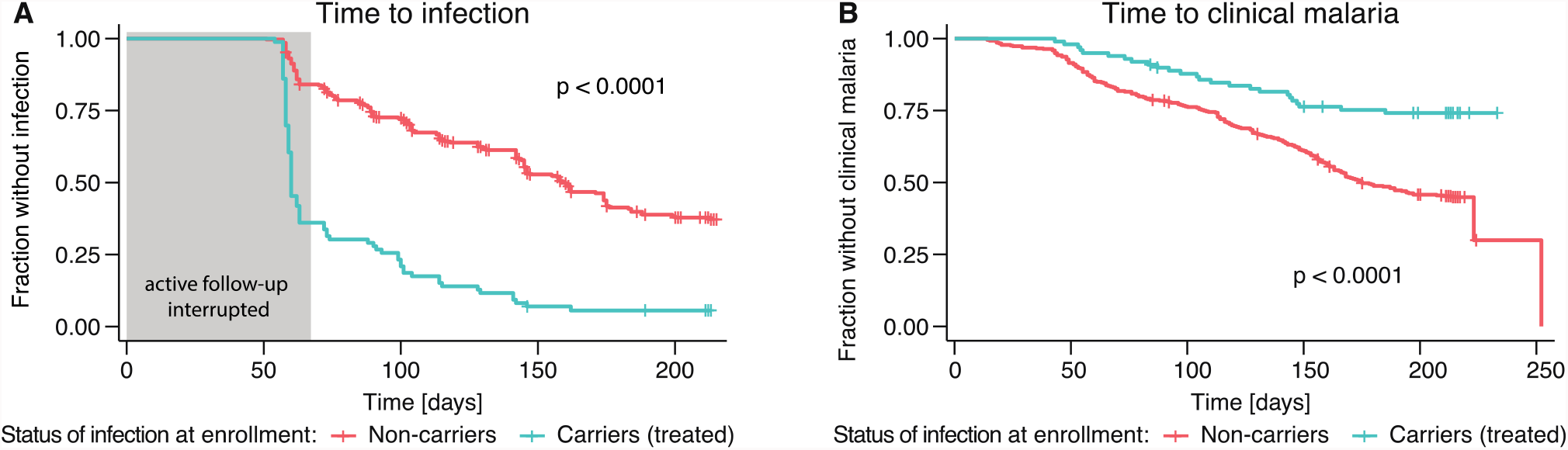
Time to infection and time to clinical malaria. (**A**) Time from enrolment to first detection of parasites with PCR for treated carriers and non-carriers. Note that active follow-up was interrupted for about 60 days between enrolment in May and the first follow-up visit in July due to limited access (see Methods and Supplementary information). Carriers have a higher risk of infection compared to non-carriers (log-rank test with p-value < 0.0001). (**B**) Time from enrolment to clinical malaria for carriers and non-carriers. This plot is a reproduction of data published by Portugal et al. (Portugal et al., 2017) who have shown that carriers are more immune to clinical malaria than non-carriers. Non-carriers have a significantly higher risk of clinical malaria than carriers (log-rank test with p-value < 0.0001). For the time to infection and clinical malaria from the first follow-up visit instead of enrolment, see ***Supplementary* Fig. S16**.

### Heterogeneity in infection risk predicted to facilitate the fast spread of malaria at the beginning of the wet season

From the dataset from Mali, it is evident that carriers have a 3.8-times higher risk of infection in the ensuing wet season compared with non-carriers (***Supplementary Fig. S13***). Carriers only account for 17.3% of the population at the end of the dry season in this data set (***Figure 6***), and here we consider whether this small subgroup of carriers allow for rapid transmission in the subsequent wet-season (Cooper et al., 2019). If mosquitos feed on dry season carriers 3.8 times as much as on non-carriers (***Supplementary Fig. S13***), then we estimate that the 17.3% of individuals who are carriers received 44.1% of all mosquito bites at the start of the wet season, due to their higher exposure (see Supplementary methods). In contrast, if all individuals were randomly received bites from the mosquito population, the dry season carrier population is expected to only receive 17.3% of all mosquito bites. In a simple model of the spread of parasites in the mosquito population, we estimate that the time until half of all the mosquito population is infected is 60.8% shorter in the case that dry season carriers are 3.8-times more likely to be bitten, compared with a scenario where bitting is homogeneous across the population (***Figure 8***). Thus, we predict that due to heterogeneity in infection risk, parasites spread 2.6 times faster in the mosquito population at the beginning of the wet season as they would if carriers and non-carriers had the same risk of infection.

**Figure 8.**
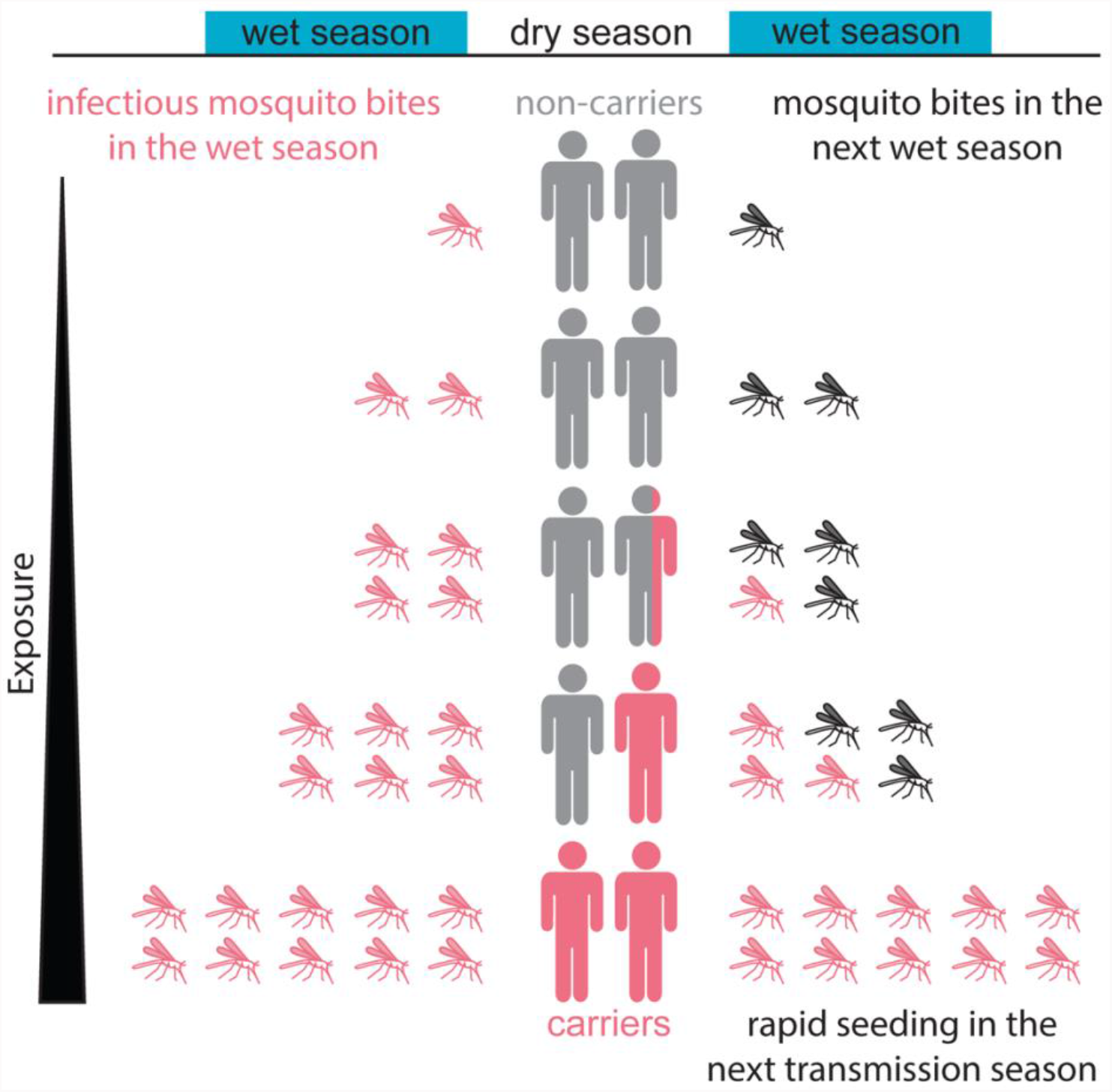
Heterogeneity in exposure facilitates the fast spread of parasites in the next transmission season. The most exposed part of the population is most likely to carry parasites over the dry season (red individuals). In the next wet season, these most-exposed individuals receive more bites than non-carriers leading to the rapid infection of the mosquito population (red mosquitoes).

## Discussion

Consistent with our proposed model where higher parasite exposure leads to an increase in duration of infections via cross-reactive host immunity, favouring dry season persistence of infections, we show data from a Malian cohort where dry season carriers have a higher risk of infection, despite their known increased protection from clinical malaria (Crompton et al., 2008; Doumbo et al., 2014; Portugal et al., 2017). These findings are also consistent with other studies that have found that in high transmission regions, immunity increases with age and the duration of infections increases with immunity (Douglas et al., 2011; Felger et al., 2012; Recker & Gupta, 2006; Rodriguez-Barraquer et al., 2018; Smith & Vounatsou, 2003). Other studies have identified exposure to infection as an important factor influencing immunity and asymptomatic parasitemia in children (Doolan et al., 2009; Wamae et al., 2019). Together these studies suggest that an individual’s history of exposure to infection is a strong determinant of the duration of their infections and parasite carriage over the dry season (***Supplementary Fig. S14***). Our modelling has shown that exposure mediated carriage of parasites leads to a seemingly contradictory observation that carriers, despite their higher protection from clinical disease, have detectable parasite loads earlier than non-carriers. This emerges because high exposure leads to increased protection from high parasite loads but not to sterilising immunity.

We found that for each level of exposure and age, there is a certain risk of persistent infections that last the entire dry season. However, there is also stochasticity in parasite carriage. It is possible that there is a “sweet spot” of immunity for parasite carriage that depends on the regularity and temporal pattern of immunity boosting infections. Indeed, it has been observed that older children (aged 5-15 years) rather than young children (less than 5 years old) or teenagers and adults (at least 16 years old) contribute disproportionately to the infectious reservoir (Andolina et al., 2021). Thus, parasite carriage may be a transient phenotype with older children being the most likely carriers since they are most likely to have had a certain level of immunity-boosting cumulative exposure without being sufficiently immune to clear all parasites during the dry season.

It has been previously observed that both super-spreading and sub-patent parasite loads play an important role in malaria transmission, in particular in low-transmission settings with malaria transmission throughout the year (Cooper et al., 2019; Slater et al., 2019). Our analysis indicates that the same is likely to be true in seasonal malaria transmission settings (Andolina et al., 2021). Carriers have a shorter time to the next infection at the beginning transmission season as well as from the first follow-up visit (about one month into the transmission season, see ***Supplementary Fig. S16***), indicating a higher level of exposure of carriers to mosquito feeding, at least early in the transmission season. However, biting rate is not the only factor determining transmission, and an individual’s potential to infect a mosquito is likely to depend on the number of gametocytes the individual carries and their infectiousness, which we have not confirmed. Although we do not have direct data on the rate at which carriers contribute to new infections of mosquitoes, our modelling suggests that heterogeneity in the biting rate with carriers being more exposed than non-carriers at the start of the transmission season may facilitate the fast spread of parasites at the beginning of the transmission season (***Figure 8***). Mosquitoes preferentially biting infected individuals (De Moraes et al., 2014; Lacroix, Mukabana, Gouagna, & Koella, 2005) may further aid in the fast spread of malaria at the beginning of the wet season. We predict that, to the extent that it is possible to identify these individuals, elimination efforts must target the most highly exposed individuals, if they aim to interrupt the emergence of parasites in the wet season.

If only a fraction of the population carries parasites over the dry season and re-seeds the following transmission season, then the question arises whether this is an advantage for malaria elimination efforts. On the one hand, fewer individuals need to be treated before the wet season as opposed to a larger proportion of the population during the wet season (Selvaraj et al., 2018). On the other hand, the difficulty lies in identifying these carriers (Stresman, Bousema, & Cook, 2019). Our modelling indicates that carriage of parasites over the dry season relates to the risk of infection but there is also stochasticity in parasite carriage since for any given FOI there is still a proportion of individuals who will not carry parasites over the dry season. Mass Drug Administration (MDA) could be a possible solution to deal with this difficulty. However, if carriers are missed in MDA programs, the effect of MDA may be short-lived due to the fast spread of parasites at the beginning of the wet season – as was suggested by previous modelling work (Kern et al., 2011; Okell et al., 2011). Repeated rounds of MDA – timed to take optimal advantage of the seasonality – in combination with vector-control may still be necessary to reduce the disease burden (Gao et al., 2020; Kern et al., 2011; Mwesigwa et al., 2019; Okell et al., 2011; Selvaraj et al., 2018).

## Materials and Methods

### Model

We adapted a model for stochastic mosquito bites and deterministic within-host dynamics (Pinkevych et al., 2012). In the model, infectious mosquito bites occur randomly. We used a piecewise constant biting rate with no bites during the dry season and a constant biting rate throughout the malaria transmission season from July through December. We incorporated parasite diversity by assuming that with every infectious mosquito bite a new parasite strain is inoculated. After the infectious bite and the liver stage of infection (which was modelled as a fixed time delay between the bite and the beginning of the blood-stage), a fixed number of parasites are released into the blood, i.e., the initial parasite concentration depends on the blood volume. Each bite induces strain specific and cross-reactive immunity. The within-host dynamics of infections are deterministic and described by the following model:

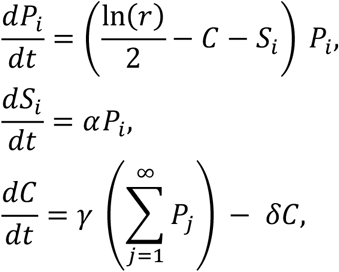

where *P*_*i*_, *i* = 1,2,3, …, is the parasite concentration of the *i*th strain (in parasites/µl), *S*_*i*_ is the strain specific immunity, and *C* is cross-reactive immunity. Thus, parasites grow at a certain rate (*r* is the PMR hence the growth rate is ln(*r*)/2), this growth is slowed by cross-reactive and strain specific immunity. If the parasite concentration falls below the clearance threshold (*Z*_*P*_), then the parasite is cleared. Strain specific immunity increases at rate *α* proportional to the parasite concentration and does not decrease in the presence of the parasite. Cross-reactive immunity increases at rate *γ* proportional to the overall parasite concentration and decreases linearly. For the interpretation of the parameters and the values used for the simulation see ***Supplementary Table S1***.

### Model simulations

The model was simulated in MATLAB (MATLAB R2018b (version 9.5.0.944444), 2018) using the parameter values in ***Table S1***. We simulated individuals with random dates of birth, random age at the beginning of the wet season (uniformly distributed between 3 months and 12 years), and a randomly chosen FOI (uniformly distributed between 0.04 and 0.004 bites per day during the dry season). If the parasite concentration of any strain decreased below the threshold *Z*_*P*_, then the parasite concentration was set to zero (without this threshold the parasite would never be cleared completely). Furthermore, the strain specific immunity of a strain that was cleared was also set to zero as it does not contribute to the within-host dynamics anymore (every infection is an infection with a new strain, the same strain is never inoculated again). At the end of the dry season, all individuals were classified as either RDT^+^, RDT^+^PCR^-^, or PCR^−^ depending on whether their overall parasite concentration was above or below the limit of detection for RDT or PCR. Treatment of RDT^+^ individuals was simulated by setting the parasite concentration to zero. All individuals were then simulated for one year. For more details on the model simulations see the Supplementary Information.

### Data from Mali

We analysed data from a longitudinal study conducted in Mali in 2012. This data set and the details of this study were previously described (Portugal et al., 2017; Tran et al., 2013). Individuals aged 3 months to 12 years were enrolled in May 2012. In the study area in Mali, malaria is seasonal with no or only very little transmission during the dry season from January through June and malaria transmission during the wet season from July through December (Portugal et al., 2017). At enrolment, i.e. at the end of the dry season, study participants were tested for *Plasmodium falciparum* parasites using a Rapid Diagnostic Test (RDT) and Principal Chain Reaction (PCR). Out of the 579 study participants, 100 (17.3%) had positive RDT results and were treated with antimalarials, 55 (9.5%) had negative RDT results but positive PCR results, and 424 (73.2%) had negative RDT and PCR results (***Figure 6***). Overall, 26.8% of children were found to carry parasites at enrolment. There is a gap of two months between enrolment (in May) and the first follow-up visit (in July) where active follow-up visits were not possible due to limited access to the study area. From July 2012 to the end of December 2012, there were 12 fortnightly follow-ups with PCR test for RDT positive (treated) and PCR negative study participants. Additionally, episodes of clinical malaria were detected by weekly active clinical surveillance visits and self-referral. The ethics committee of the Faculty of Medicine, Pharmacy and Dentistry at the University of Sciences, Techniques and Technology of Bamako, and the Institutional Review Board of NIAID NIH approved the study (ClinicalTrials.gov NCT01322581). Written, informed consent was obtained from the parents or guardians of participating children or from adult participants.

### Data analysis and statistical information

The software R (version 3.6.0) (R Core Team, 2019) was used to analyse the data. The data from Mali was analysed using the package dplyr (version 0.8.3) (Wickham, François, Henry, & Müller, 2019) and survival analysis with the packages survival (version 2.38) (T. Therneau, 2015; T. M. Therneau & Grambsch, 2000) and survminer (version 0.4.5) (Kassambara, Kosinski, & Biecek, 2019). In boxplots, the length of the whiskers is the minimum of 1.5 times the interquartile range and the distance from the box to the largest or lowest (for upper and lower whiskers respectively) data value. Survival curves were compared with the log-rank test and different groups were compared with one-sided Wilcoxon rank-sum tests. We excluded duplicates and data for the same individual (same ID) but other conflicting values. If there were inconsistencies in the follow-up dates (e.g., third visit before the second visit), then we only considered the follow-ups before the first inconsistent date. For time from first follow-up to PCR^+^, we excluded individuals with a positive PCR result at the first follow-up visit and we allowed maximally 20 days between known PCR results. If the time between two known PCR results is more than 20 days, then we consider this case as right censored before this gap. We considered the time from first follow-up instead of time from enrolment to first positive due to the gap of approximately 60 days between enrolment and the first follow-up with a PCR test (we obtained the same results for considering the time from enrolment and the time from the first follow-up visit, see Supplementary results). For time to clinical malaria, if an individual had no observed episode of clinical malaria, then we considered this individual as right censored at the date of the last follow-up.

## Supporting information

Supplementary Infiormation

## Data Availability

The data and code are available upon request.

## Data Availability

The data and code are available upon request.

## Acknowledgements

This work was funded by the Australian Research Council (ARC) (grant DP120100064 & DP180103875 (to DSK, MPD) and the National Health and Medical Research Council (NHMRC) of Australia (grants 1082022 (to MPD), 1141921 (to DSK), and 1080001 and 1173027 (to MPD). The cohort study in Mali was funded by the Division of Intramural Research, National Institute of Allergy and Infectious Diseases, National Institutes of Health, with intramural grants to PDC.

## Author contributions

ES, DC, SO, PDC, SP, MPD and DSK contributed to modelling, simulations and statistical analysis. AO, SD, KK, BT, PDC and SP contributed to data collection and curation. ES, DC, PDC, SP, MPD and DSK contributed to writing the manuscript and visualisations. All authors reviewed and approved the final manuscript and had final responsibility for the decision to submit for publication.

## Data and code availability

The data and code are available upon request.

