## Supplementary Infiormation for "A role for super-spreaders in carrying malaria parasites across the months-long dry season"

**Supplementary information**

Eva Stadler[1], Deborah Cromer[1], Samson Ogunlade[1], Aissata Ongoiba[2], Safiatou Doumbo[2], Kassoum Kayentao[2], Boubacar Traore[2], Peter D Crompton[3], Silvia Portugal[3], Miles P Davenport[1], David S Khoury[1]*

[1] The Kirby Institute, UNSW Sydney, Sydney, NSW 2052, Australia
[2] Malaria Research and Training Centre, Department of Epidemiology of Parasitic Diseases, International Center of Excellence in Research, University of Sciences, Technique, and Technology of Bamako, 91094, Bamako, Mali
[3] Malaria Infection Biology and Immunity Section, Laboratory of Immunogenetics, National Institute of Allergy and Infectious Diseases, National Institutes of Health, Rockville, USA

### Supplementary tables

**Parameters of the model**

| Parameter | Interpretation | Value [unit] | Reference |
|---|---|---|---|
| r | Initial Parasite Multiplication Rate (PMR) | 14 | [1-3] |
| $\alpha$ | Rate of increase of strain specific immunity | $7 \times 10^{-6}$ [$\mu$L / (par. × day)] | [a] |
| $\gamma$ | Rate of increase of cross-reactive immunity | $1 \times 10^{-6}$ [$\mu$L / (par. × day)] | [b] |
| $\delta$ | Rate of loss of cross-reactive immunity | $3.7 \times 10^{-4}$ [day$^{-1}$] | [4, 5] |
| $\varepsilon$ | Initial number of infected RBCs | 56,000 [par.] | [1] |
| lod | Limit of detection for PCR | 1 [par./$\mu$L] | [6, 7] |
| lod_rdt | Limit of detection for RDT | 100 [par./$\mu$L] | [6] |
| $Z_P$ | Threshold for parasite growth | $2 \times 10^{-7}$ [par./$\mu$L] | [1] |
| V(t) | Blood volume (in liters) depending on time since birth (in days) | $V(t) = \begin{cases} 0.00059\, t + 0.3, & \text{if } t < 22 \times 365, \\ 5, & \text{if } t \geq 22 \times 365. \end{cases}$ | [1] |
| | Duration of the liver-stage | 7 [days] | [8] |
| | Biting rate during the dry season (i.e., days 1 to 181) | 0 [bites/day] | |
| | Biting rate during the malaria transmission season (i.e., days 182 to 365) | 1/250 – 1/25 [bites/day] | [b] |

**Table S1** Parameters of the model, their interpretation, value, unit, and reference.

[a] The growth of strain specific immunity, $\alpha$, is chosen such that the maximal parasite concentration during the first infection is approximately $10^5$ parasites/$\mu$L (the parasite concentration at first infection in naïve neurosyphilis patients[9-11]). Thus, for $r = 14$ we choose $\alpha = 7 \times 10^{-6}$.

[b] The rate of increase of cross-reactive immunity, $\gamma$, and the biting rate during the malaria transmission season were chosen together such that the fraction of PCR⁻ individuals in the heterogeneous simulation is close to the fraction of PCR⁻ individuals in the data (in the data 73% of the individuals are PCR⁻).

**Overview over simulations of homogeneous populations with the same FOI**

| FOI [bites per day] | Number of carriers | Time to first bite (p-value) | Time to PCR positive (p-value) | Older (p-value) | Lower PMR (p-value) | Higher cross-reactive immunity (p-value) |
|---|---|---|---|---|---|---|
| 0.004 | 1 | n.s. (0.84) | n.s. (0.2) | n.s. (0.14) | carriers (0.0095) | n.s. (0.13) |
| 0.008 | 0 | - | - | - | - | - |
| 0.012 | 11 | n.s. (0.87) | n.s. (0.22) | carriers (0.013) | n.s. (0.44) | n.s. (0.46) |
| 0.016 | 54 | n.s. (0.45) | n.s. (0.22) | carriers (<0.0001) | n.s. (0.76) | carriers (0.00046) |
| 0.020 | 101 | n.s. (0.32) | different (0.038) | carriers (<0.0001) | non-carriers (0.0079) | carriers (<0.0001) |
| 0.024 | 205 | n.s. (0.42) | n.s. (0.2) | carriers (<0.0001) | n.s. (0.19) | carriers (<0.0001) |
| 0.028 | 315 | n.s. (0.079) | different (0.049) | carriers (<0.0001) | non-carriers (<0.0001) | carriers (<0.0001) |
| 0.032 | 399 | n.s. (0.33) | different (0.0055) | carriers (<0.0001) | non-carriers (<0.0001) | carriers (<0.0001) |
| 0.036 | 524 | n.s. (0.16) | different (0.015) | carriers (<0.0001) | non-carriers (<0.0001) | carriers (<0.0001) |
| 0.040 | 625 | n.s. (0.32) | n.s. (0.62) | carriers (<0.0001) | non-carriers (<0.0001) | carriers (<0.0001) |

**Table S2** Comparison of different characteristics of carriers and non-carriers in the simulations of homogeneous populations with different FOIs. Individuals were classified as carriers if they have a parasite concentration above the limit of detection for a Rapid Diagnostic Test (RDT) at the end of the dry season and as non-carriers if they had a negative PCR result at the end of the dry season. For each FOI, a population of 1,000 individuals with different age was simulated as described in the Supplementary methods "Model simulation for a heterogeneous population" with the only difference that the biting rate during the wet season was chosen to be the same for all individuals. The time to first bite, age, PMR and cross-reactive immunity of carriers and non-carriers were compared with the Wilcoxon rank-sum test (two-sided test and if significant, i.e., p-value < 0.05, then also with a one-sided Wilcoxon rank-sum test). The time to PCR positive was compared with a log-rank test. For the time to PCR positivity see **Fig. S7**. Abbreviations: FOI force of infection, n.s. not significant, PMR parasite multiplication rate.

### Supplementary figures

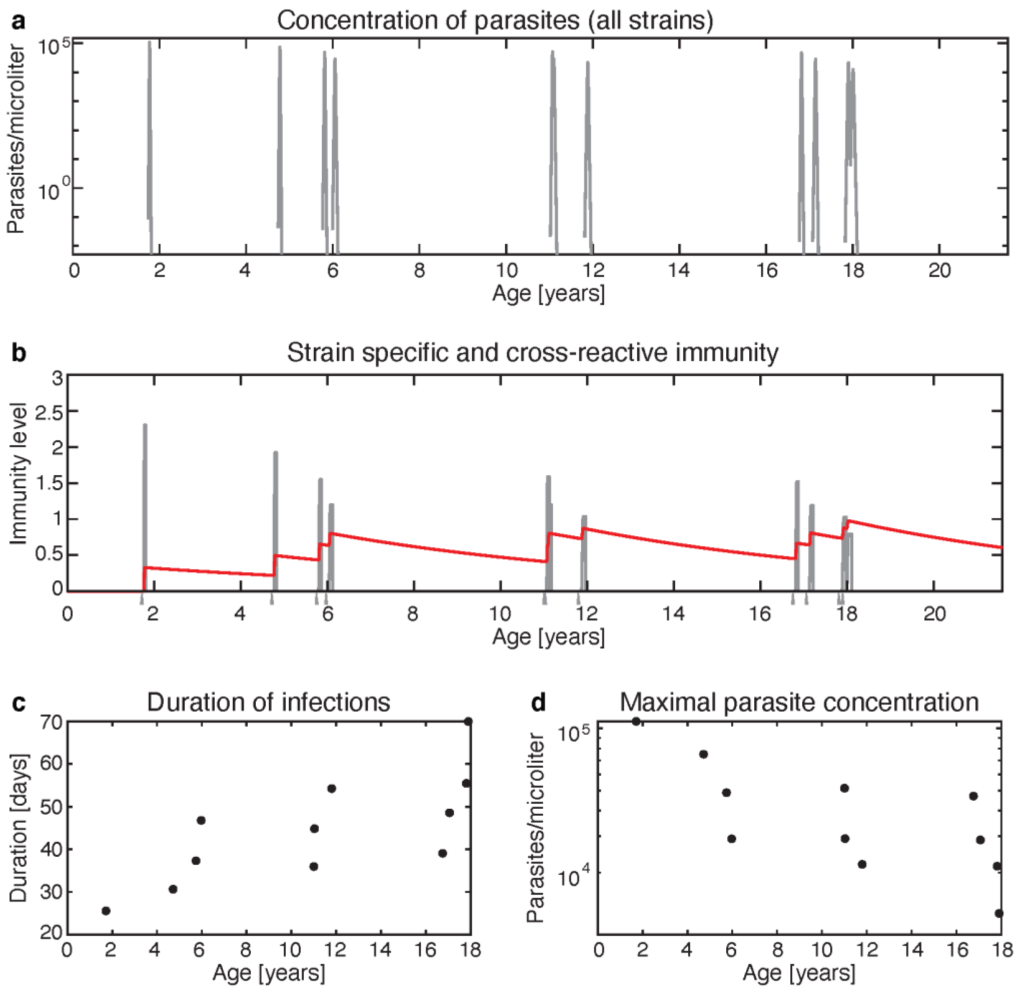


**Fig. S1** Simulation of a single individual from birth. The biting rate during the malaria transmission season was chosen as the minimal biting rate, i.e., 0.004 bites per day. All other parameters are as in **Table S1**. **a** Parasite concentration of all strains vs age. **b** Strain specific immunity (gray lines) and cross-reactive immunity (red line) vs age. Note that strain specific immunity is only plotted for the time interval the respective strain is present in the blood. If the parasite is cleared from the blood, then strain specific immunity to this strain only decreases exponentially and neither influences cross-reactive immunity nor the dynamics of infections with other strains. The gray arrows at the bottom of the plot indicate mosquito bites. **c** Duration of infections from the beginning of the blood-stage until the parasite strain is cleared. **d** Maximal parasite concentration of each infection.

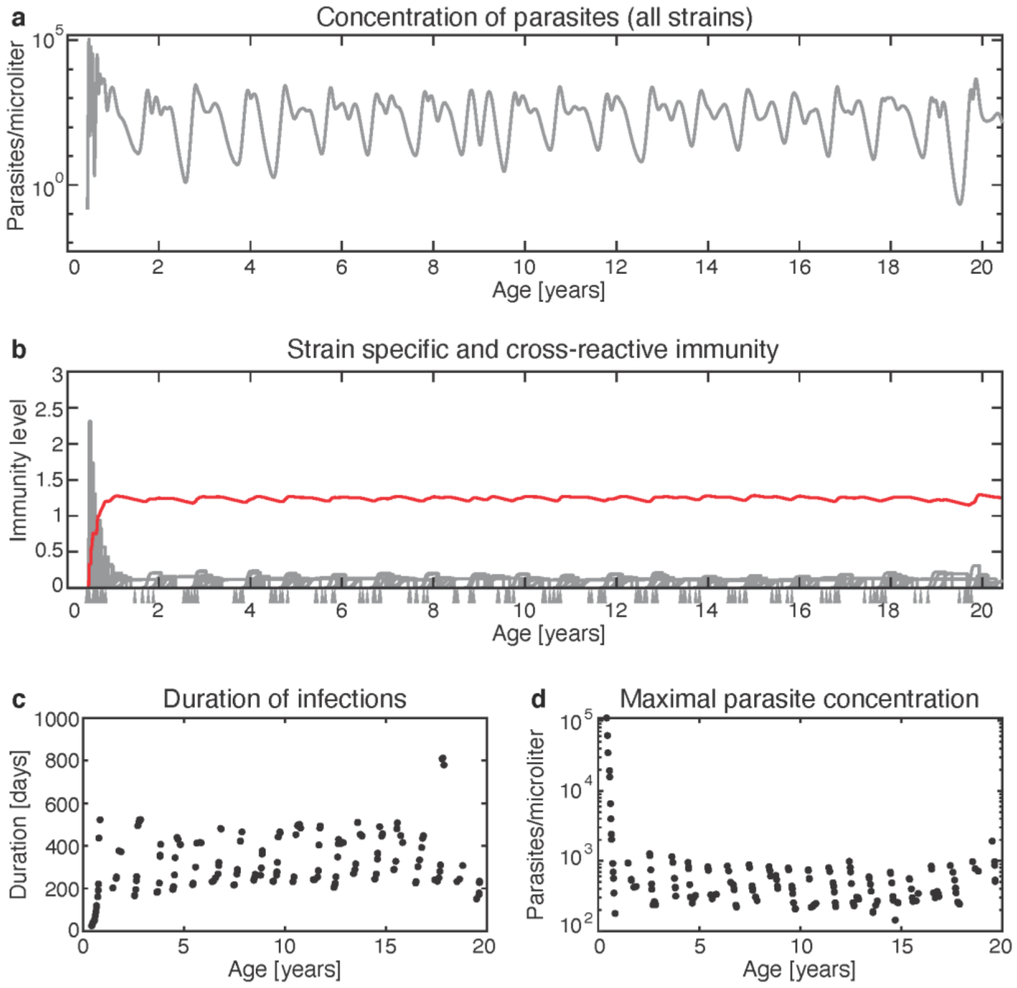


**Fig. S2** Simulation of a single individual from birth. The biting rate during the malaria transmission season was chosen as the maximal biting rate, i.e., 0.04 bites per day. All other parameters are as in **Table S1**. **a** Parasite concentration of all strains vs age. **b** Strain specific immunity (gray lines) and cross-reactive immunity (red line) vs age. Note that strain specific immunity is only plotted for the time interval the respective strain is present in the blood. If the parasite is cleared from the blood, then strain specific immunity to this strain only decreases exponentially and neither influences cross-reactive immunity nor the dynamics of infections with other strains. The gray arrows at the bottom of the plot indicate mosquito bites. **c** Duration of infections from the beginning of the blood-stage until the parasite strain is cleared. **d** Maximal parasite concentration of each infection.

Age of first parasite carriage at the end of the dry season

| Force Of Infection (FOI) [bites/day] \ Age of first parasite carriage [years] | 0-1 | 1-2 | 2-3 | 3-4 | 4-5 | 5-6 | 6-7 | 7-8 | 8-9 | 9-10 | 10-11 | 11-12 | 12-13 | 13-14 | 14-15 | 15-16 | 16-17 | 17-18 | 18-19 | 19-20 |
|---|---|---|---|---|---|---|---|---|---|---|---|---|---|---|---|---|---|---|---|---|
| 0.04 | 0.24 | 14.22 | 44.95 | 26.55 | 9.44 | 3.36 | 0.96 | 0.24 | 0 | 0.04 | 0 | 0 | 0 | 0 | 0 | 0 | 0 | 0 | 0 | 0 |
| 0.036 | 0.1 | 8.93 | 35.7 | 31.3 | 14.21 | 6.44 | 2.39 | 0.61 | 0.25 | 0.07 | 0 | 0 | 0 | 0 | 0 | 0 | 0 | 0 | 0 | 0 |
| 0.032 | 0.01 | 4.85 | 24.76 | 28.54 | 20.76 | 10.52 | 5.38 | 2.85 | 1.1 | 0.49 | 0.35 | 0.18 | 0.09 | 0.08 | 0 | 0 | 0.04 | 0 | 0 | 0 |
| 0.028 | 0 | 2.55 | 14.17 | 23.07 | 19.2 | 13.14 | 9.93 | 6.23 | 4.23 | 3.06 | 1.61 | 0.92 | 0.76 | 0.5 | 0.22 | 0.19 | 0.09 | 0.05 | 0.08 | 0 |
| 0.024 | 0 | 0.79 | 6.44 | 13.28 | 16.88 | 13.69 | 11.91 | 8.56 | 7.52 | 4.38 | 4.43 | 2.92 | 2.35 | 1.66 | 1.31 | 0.85 | 0.69 | 0.61 | 0.36 | 0.4 |
| 0.02 | 0 | 0.39 | 2.06 | 6.01 | 8.12 | 8.38 | 8.61 | 8.38 | 7.56 | 6.45 | 4.95 | 4.94 | 4.61 | 4 | 3.67 | 3.14 | 2.57 | 2.06 | 1.82 | 1.68 |
| 0.016 | 0 | 0.04 | 0.27 | 1.26 | 2.25 | 2.66 | 4.01 | 3.47 | 4.11 | 3.5 | 3.54 | 3.56 | 3.42 | 3.57 | 2.85 | 3.09 | 2.93 | 2.91 | 3.04 | 1.86 |
| 0.012 | 0 | 0 | 0.07 | 0.15 | 0.3 | 0.28 | 0.55 | 0.61 | 0.61 | 0.8 | 0.92 | 0.97 | 0.72 | 0.68 | 0.8 | 0.85 | 0.81 | 0.8 | 1.08 | 0.91 |
| 0.008 | 0 | 0 | 0 | 0 | 0.06 | 0 | 0 | 0.03 | 0.03 | 0.06 | 0 | 0.08 | 0 | 0 | 0.04 | 0.03 | 0 | 0.08 | 0.04 | 0.01 |
| 0.004 | 0 | 0 | 0 | 0 | 0 | 0 | 0 | 0 | 0 | 0 | 0 | 0 | 0 | 0 | 0 | 0 | 0 | 0 | 0 | 0 |

Colour scale: 0%, 10%, 20%, 30%, 40%, 50%, 60%, 70%, 80%, 90%, 100%

**Fig. S3** Age of first carriage of parasites over the dry season by FOI during the wet season and age. For each FOI, 10,000 individuals were simulated from birth to age 20. Individuals with a higher force of infection carry parasites at a younger age. Some individuals with a lower FOI never carry parasites over the dry season before they reach age 20.

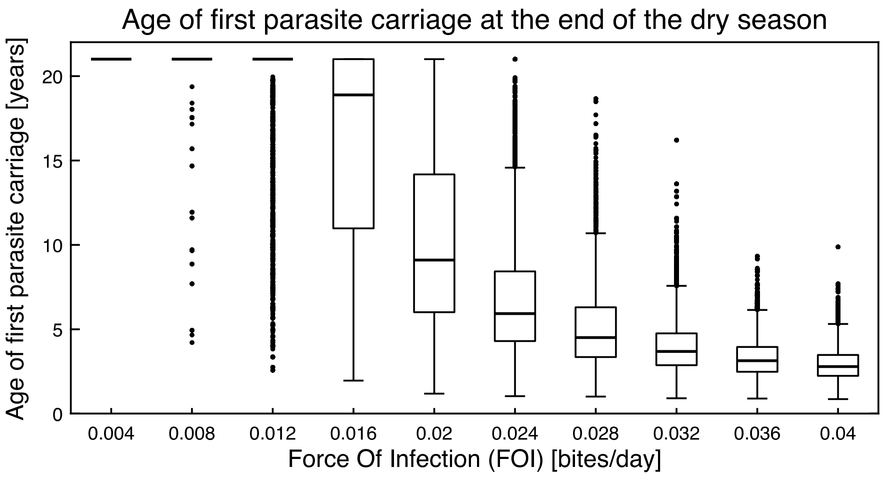


**Fig. S4** Age of first parasite carriage over the dry season by FOI. Simulation for 100,000 individuals. For each FOI 10,000 individuals were simulated from birth to age 20. Note that there is heterogeneity in the age of first carriage that is due to stochasticity in the mosquito bites. The heterogeneity in age of first carriage is greatest at intermediate FOIs.

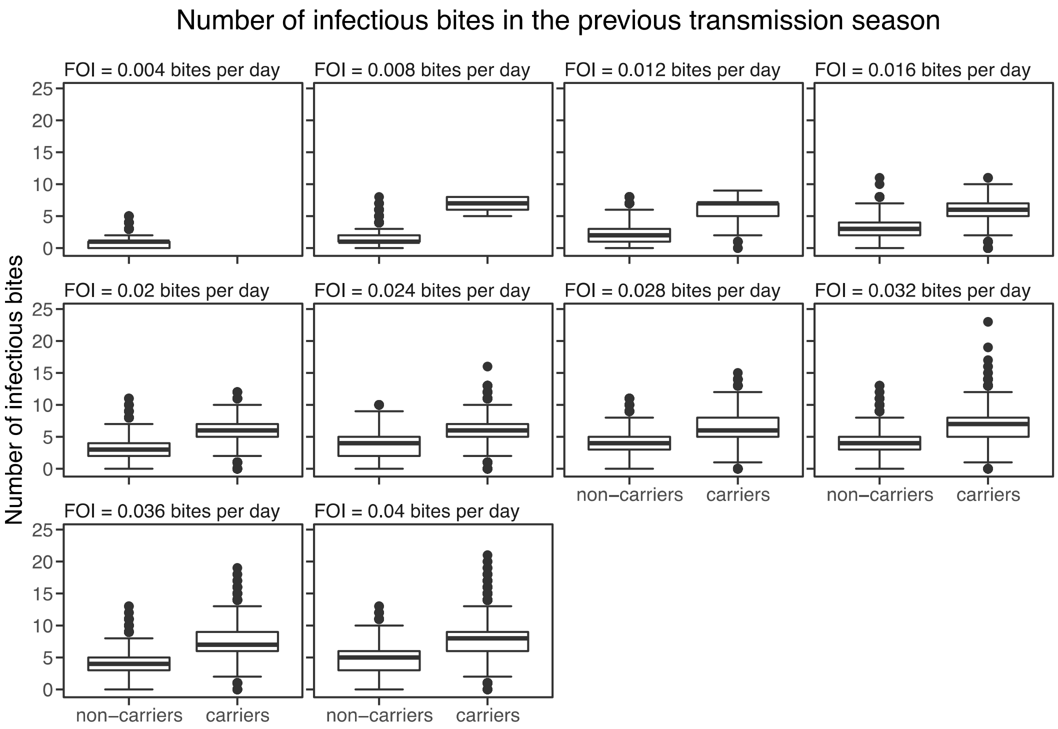


**Fig. S5** Number of infectious bites in the previous transmission season for carriers and non-carriers in the simulations with homogeneous FOI. For each FOI, 10,000 individuals were simulated from birth to age 20, then for each individual an age was chosen uniformly between 0 and 19. At the chosen age, individuals were classified as carriers if they had a parasite concentration above the limit of detection for a Rapid Diagnostic Test (RDT) at the end of the dry season. This figure shows the number of infectious bites carriers and non-carriers received in the transmission season prior to their classification as either a carrier or a non-carrier. Carriers had significantly more infectious bites in the previous season than non-carriers (one-sided Wilcoxon rank-sum test with p-value < 0.0001 for each FOI, except for 0.004 bites per day as there were no carriers for this FOI).

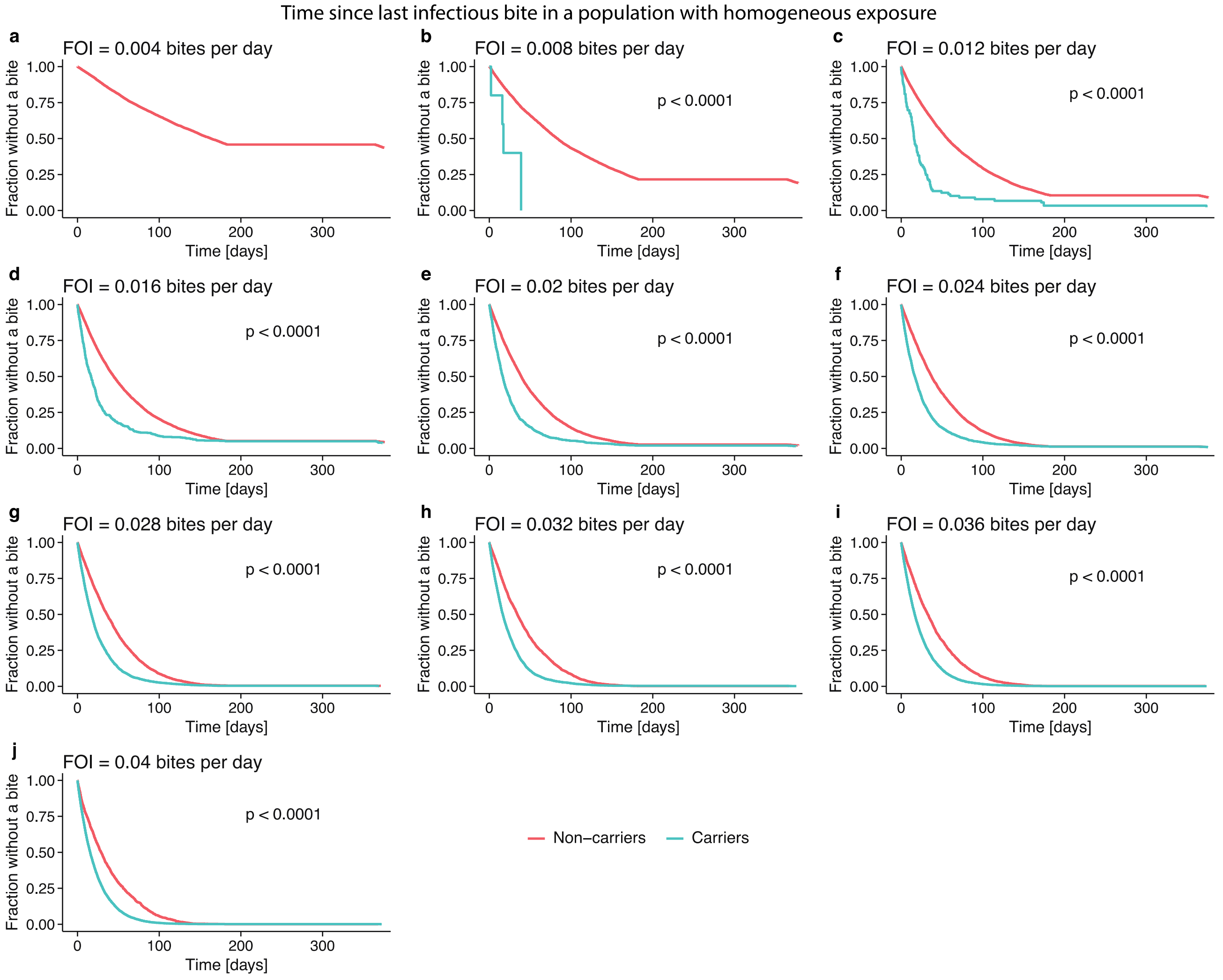


**Fig. S6** Time since last infectious bite for carriers and non-carriers in the simulations with homogeneous FOI. For each FOI, 10,000 individuals were simulated from birth to age 20, then for each individual an age was chosen uniformly between 0 and 19. At the chosen age, individuals were classified as carriers if they had a parasite concentration above the limit of detection for a Rapid Diagnostic Test (RDT) at the end of the dry season. This figure shows the time from the last infectious bite to the end of the transmission season prior to classification as a carrier or non-carrier (the time to the end of the dry season would just be an additional 181 days for all individuals). For each FOI (except 0.004 bites per day as there were no carriers for this FOI), carriers have had a significantly shorter time since the last infectious bite (log-rank test with p-value < 0.0001 for all FOIs). Thus, carriers had a more recent infectious bite than non-

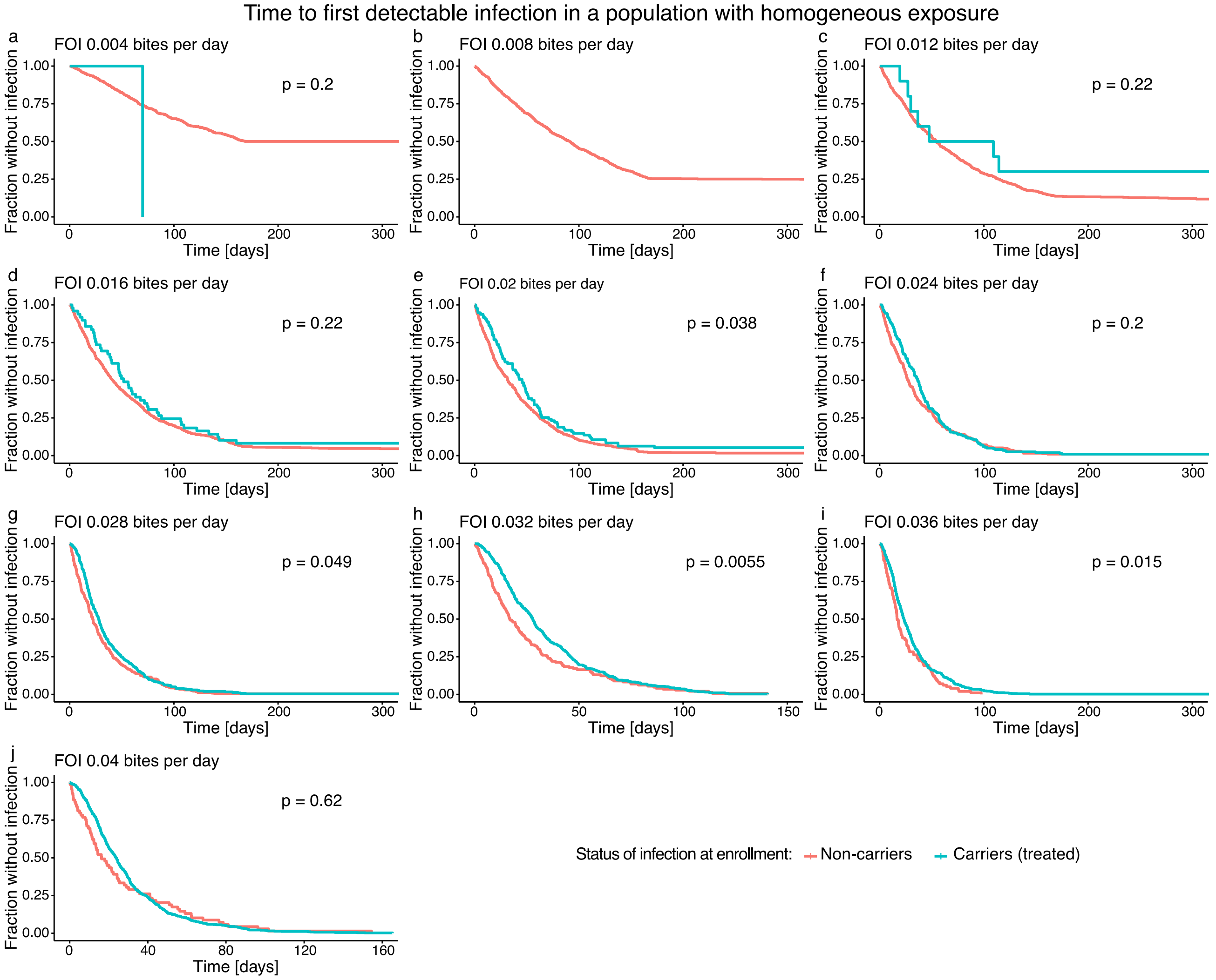


**Fig. S7** Time from first follow-up to positive PCR result for a population of 1,000 individuals with the same FOI but different age. Each subfigure shows the time to detectable infection for a different FOI. The curves are compared with the log-rank test (p-values in the upper right corner). Note that the time from first follow-up to $PCR^+$ is either not significantly different between carriers and non-carriers or appear to be longer for carriers than for non-carriers (unlike what we observe for a heterogeneous population). Abbreviations: FOI force of infection.

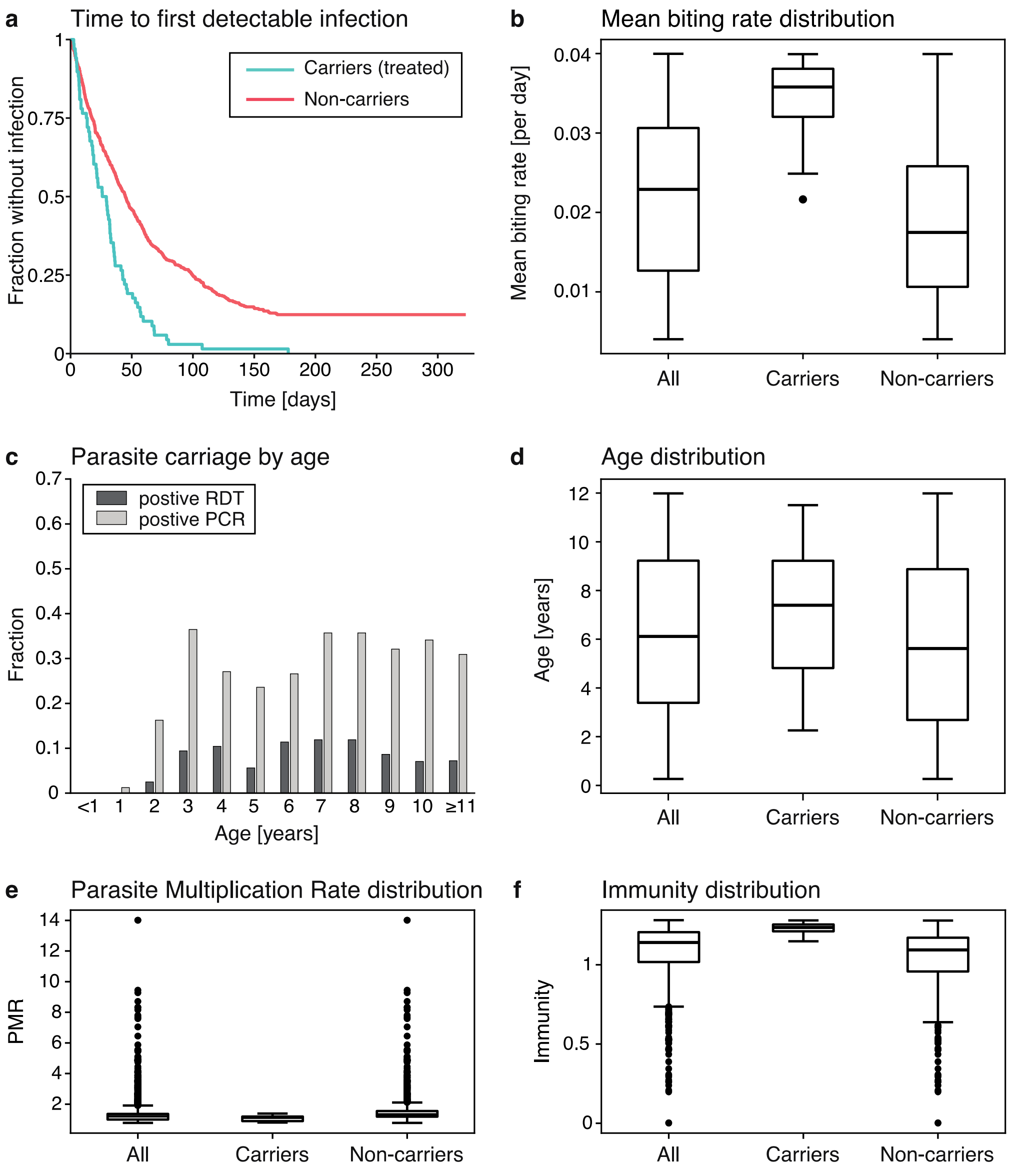


**Fig. S8** Simulation of 1000 individuals aged 3 months to 12 years with different FOIs during the wet season. Individuals were classified as carriers or non-carriers depending on whether they have a parasite concentration above the limit of detection for a Rapid Diagnostic Test (RDT) at the end of the dry season. The simulation included treatment of carriers. Overall, there were 74 RDT positive (treated), 184 RDT negative and PCR positive, and 742 PCR negative individuals at the end of the dry season. The parameter values for this simulation can be found in **Table S1**. **a** Time from the first follow-up to infection (by PCR, i.e., the overall parasite concentration exceeds the limit of detection for PCR). Carriers had a significantly higher infection risk compared to non-carriers (log-rank test with p-value < 0.0001). **b** FOI for all individuals, carriers, and non-carriers. Carriers have a significantly higher mean biting rate than non-carriers (Wilcoxon rank-sum test with p-value < 0.0001). **c** Fraction of individuals that are RDT positive and PCR positive by age. A higher fraction of the older children carries parasites at the end of the dry season compared to younger children. **d** Age distribution of all individuals, carriers, and non-carriers. Carriers are significantly older than non-carriers (Wilcoxon rank-sum test with p-value 0.0005). **e** Parasite Multiplication Rate (PMR) distribution of all individuals, carriers, and non-carriers for each strain at the end of the surveillance period. The PMRs of carriers are significantly lower than those of non-carriers (Wilcoxon rank-sum test with p-value < 0.0001). **f** Cross-reactive immunity distribution of all individuals, carriers, and non-carriers. Carriers have a significantly higher cross-reactive immunity than non-carriers (Wilcoxon rank-sum test with p-value < 0.0001).

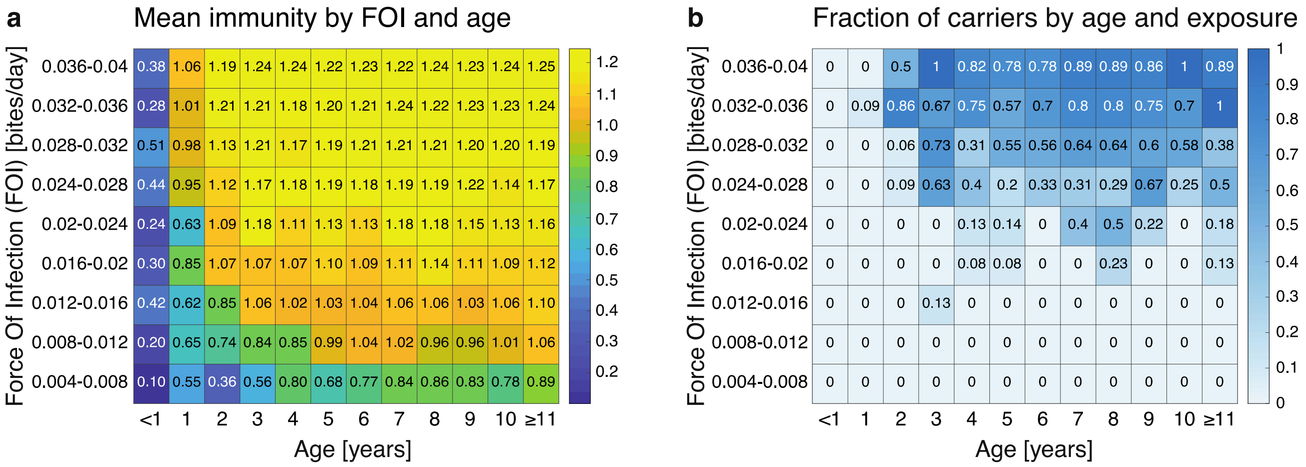


**Fig. S9** Immunity and parasite carriage by age and FOI in the simulation of 1,000 individuals with heterogeneous age and FOI. **a** Mean cross-reactive immunity by FOI and age. Children who are older and have a higher FOI also have higher mean cross-reactive immunity. **b** Fraction of carriers by FOI and age. Here, carriers are children who have a parasite concentration above the limit of detection of PCR at the end of the dry season. The highest fractions of carriers can be found among children who are older and have a higher FOI.

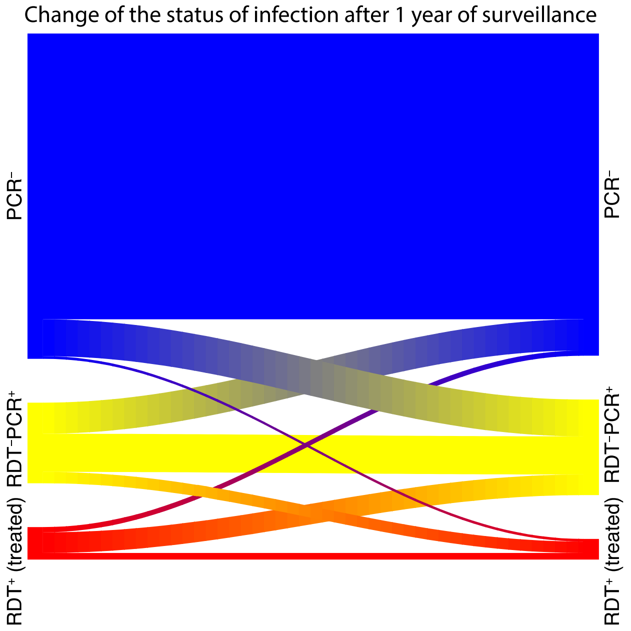


**Fig. S10** Change of the status of infection in the model simulation of 1,000 individuals after 1 year of surveillance. On the left-hand side is the status of infection at the beginning of the simulation and on the right-hand side is the status of infection after a simulated surveillance period of 1 year. Overall, there were 1,000 individuals. Initially there are 742 individuals with negative PCR result ($PCR^-$), after 1 year, 6 of them are $RDT^+$ individuals (positive Rapid Diagnostic Test), 84 are $RDT^-PCR^+$ (negative Rapid Diagnostic Test but positive PCR result), and 652 are still $PCR^-$. Of 184 $RDT^-PCR^+$ individuals 26 become $RDT^+$, 87 remain $RDT^-PCR^+$, and 71 become $PCR^-$. Of 74 $RDT^+$ individuals 15 remain $RDT^+$, 47 become $RDT^-PCR^+$, and 12 become $PCR^-$. After 1 year there are overall 47 $RDT^+$, 218 $RDT^-PCR^+$, and 735 $PCR^-$.

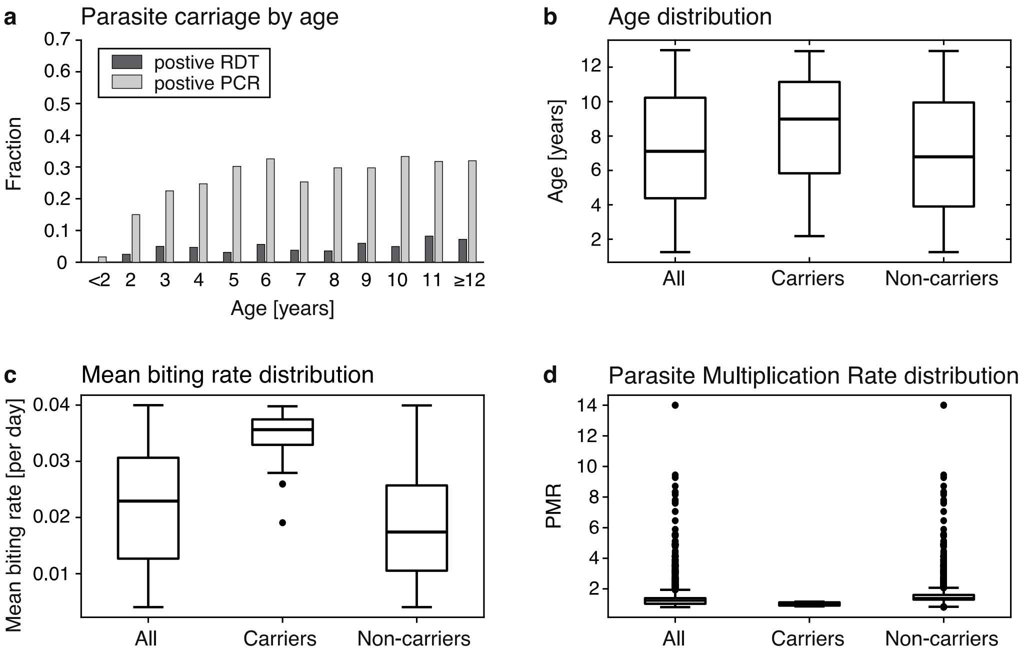


**Fig. S11** Characterization of carriers after one year of surveillance in the model simulations. **a** Fraction of RDT⁺ and PCR⁺ for different ages. **b** Age distribution of carriers and non-carriers. Carriers are significantly older than non-carriers (Wilcoxon rank-sum test with p-value 0.005). **c** Mean biting rate for carriers and non-carriers. Carriers have a significantly higher mean biting rate than non-carriers (Wilcoxon rank-sum test with p-value < 0.0001). **d** Parasite Multiplication Rate (PMR) of carriers and non-carriers. Carriers have a significantly lower PMR than non-carriers (Wilcoxon rank-sum test with p-value < 0.0001).

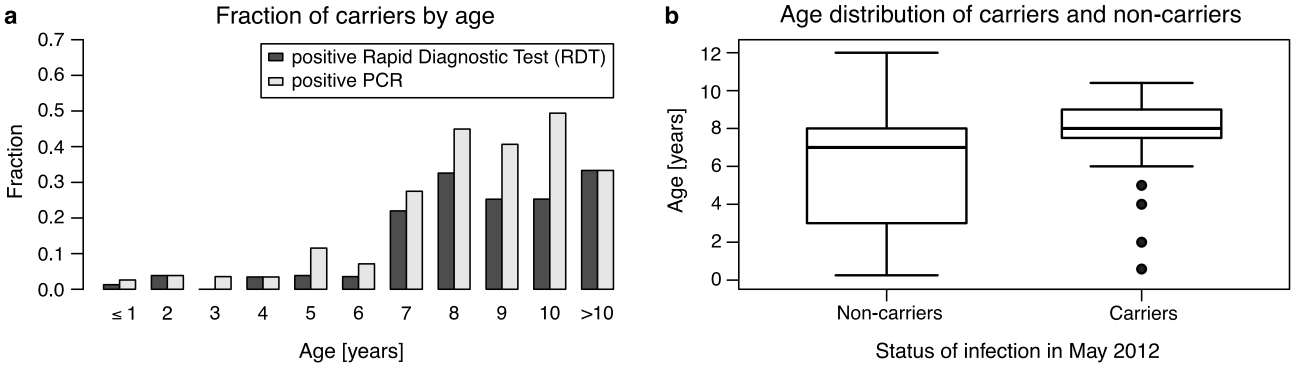


**Fig. S12** Age and status of infection at the end of the dry season in the data. **a** Fraction of individuals with a positive Rapid Diagnostic Test (RDT⁺) and positive PCR result (PCR⁺) at enrolment, i.e. at the end of the dry season, by age. **b** Age distribution of non-carriers and carriers. Carriers are significantly older than non-carriers (Wilcoxon rank-sum test with p-value < 0.0001).

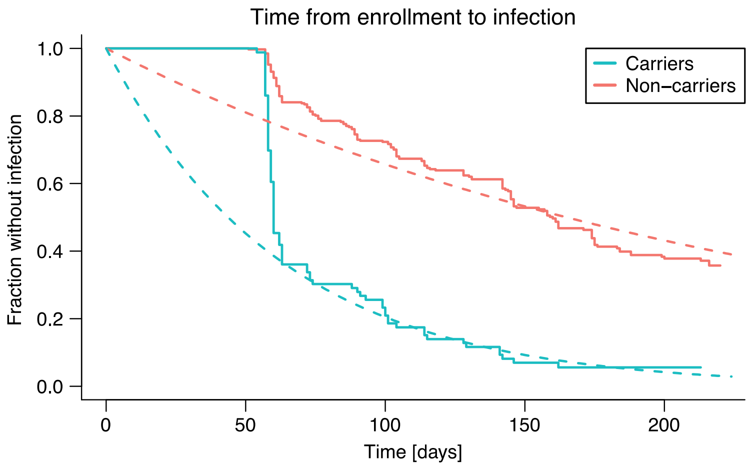


**Fig. S13** Fit of a constant biting rate to the time from enrollment to infection in the data, with left censoring for the first 70 days after enrollment. The fit is shown as a dashed line. The estimated biting rate for carriers is 0.016 bites per day and the estimated biting rate for non-carriers is 0.0042 bites per day.

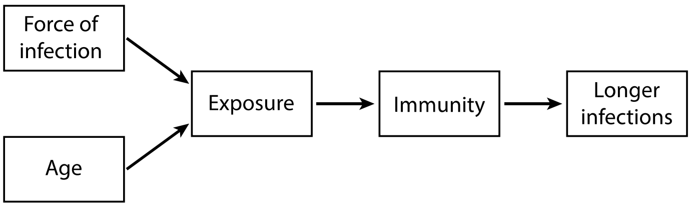


**Fig. S14** Schematic of the model-based hypothesis from the simulation of a heterogeneous population. With age and FOI, the (cumulative) exposure increases. Exposure leads to higher levels of immunity which in turn leads to longer infections and carriage of parasites through the dry season.

### Supplementary methods

**Model simulation for a single individual**
An individual was simulated by randomly choosing a time in the year as the time of birth. Initially, the individual has no strain specific or cross-reactive immunity and is parasite-free. The time to the first infectious mosquito bite and the time between two mosquito bites was computed using inverse transform sampling (see below for the details). The biting rate during the dry season is zero for all individuals, the mean biting rate during the malaria transmission was chosen randomly (uniformly) in the range specified in **Table S1**. The mean biting rate for an individual during the malaria transmission season is the same every year, it does not vary with time. Each infectious bite is an infection with a new parasite strain.
The deterministic within host dynamics (see main text for the model equations) were simulated in MATLAB (R2018b)[12] using the ordinary differential equations solver ode45. If the parasite concentration dropped below the threshold $Z_P$, then the parasite was cleared, i.e., the concentration was set to 0. As each infection is an infection with a new strain, the strain specific immunity of a strain that was cleared does not contribute to the within-host dynamics anymore and was thus also set to zero.
Simulations for single individuals with the minimal and the maximal mosquito biting rates are shown in **Fig. S1** and **Fig. S2**, respectively. If an individual does not receive new infections anymore and had a low Force Of Infection (FOI) before infectious bites stopped, then parasites are cleared and cross-reactive immunity decreases (see **Fig. S1**).

**Time to the next infectious mosquito bite**
The time to the first infectious mosquito bite and the time between two mosquito bites was computed using a non-homogeneous Poisson process with a piecewise constant biting rate (the biting rate is constant on each day). The biting rate during the dry season is zero for all individuals, during the wet season it is constant for each individual but varies between 0.004 and 0.04 bites per day for different individuals (**Table S1**).
In order to compute the time to the next bite, we use inverse transform sampling, i.e., we sample a number $x$ from a uniform distribution between 0 and 1 and determine the time $t$ such that the $F^{-1}(x) = t$, where $F(t)$ is the cumulative distribution function (cdf) for the time to the next bite. Since the biting rate is constant for each day, the time to the next bite is exponentially distributed and the cdf for the time to the next bite is given by

$$F(t) = 1 - e^{-\int_a^t b(y)\,dy},$$

where $b(y)$ is the biting rate at time $y$, $a$ is the current time, and $t$ is the time to the next bite. If $F^{-1}(x) = t$ or $F(t) = x$ for $x$ sampled from a uniform distribution between 0 and 1, then

$$\int_a^t b(y)\,dy = -\log(1 - x).$$

Using that the biting rate $b(y)$ is constant each day this expression simplifies to

$$(\lceil a \rceil - a) \times b(a) + \sum_{i=1}^{n} b(a + i) + (t - \lfloor t \rfloor) \times b(t) = -\log(1 - x),$$

where $n$ is the number of days such that

$$(\lceil a \rceil - a) \times b(a) + \sum_{i=1}^{n} b(a+i) \leq -\log(1-x)$$

and

$$(\lceil a \rceil - a) \times b(a) + \sum_{i=1}^{n+1} b(a+i) > -\log(1-x).$$

With $n$ computed in this way, we compute $d = t - \lfloor t \rfloor$ in the following way:

$$d = \frac{-\log(1-x) - (\lceil a \rceil - a) \times b(a) - \sum_{i=1}^{n} b(a+i)}{b(a+n+1)}.$$

Overall, the time to the next bite is then given by

$$t = (\lceil a \rceil - a) + n + d.$$

**Age of first parasite carriage**

We simulated individuals as described above and computed for each individual the age of first carriage of parasites over the dry season. An individual was considered to carry parasites over the dry season if the overall parasite concentration on the last day of the dry season was above the limit of detection of PCR. The result of the simulation is shown in **Fig. 3**, **Fig. S3**, and **Fig. S4**.

**Model simulation for a heterogeneous population**

We simulated 1,000 individuals. Each individual was assigned a random (uniformly chosen) age at the end of the dry season that we chose as June 30th. Then, all individuals were simulated from birth to their age at the end of the dry season as explained above (see Model simulation for a single individual) to determine their parasite density, strain-specific immunity for each strain they carry, and level of cross-reactive immunity at the beginning of the one-year follow-up. At the end of the dry season, they were classified as $RDT^+$, $RDT^+PCR^-$, or $PCR^-$ depending on their parasite concentration. Treatment of $RDT^+$ individuals was simulated by setting the parasite concentration and strain-specific immunity to zero for all strains. Then, all individuals were simulated for a one-year surveillance period.

The Parasite Multiplication Rate (PMR) of strain $i$ at time $t$ was computed as follows:

$$\mathrm{PMR}_i(t) = r \times e^{-2(S_i(t)+C(t))},$$

where $S_i(t)$ is the strain specific immunity and $C(t)$ is the cross-reactive immunity at time $t$. In **Fig. S8** and **Fig. S11**, we report the PMR for each individual and each strain at the end of the surveillance period, i.e., at the end of the dry season one year after enrollment.

In the model simulations, we can observe how the status of infection changes over the duration of the one-year surveillance period (**Fig. S10**). Even though the status of infection changes for 246 out of 1000 individuals, we still find that carriers are significantly older, more exposed, and more immune than non-carriers if we consider carriers at the end of the surveillance time instead of carriers at the beginning of the surveillance time (**Fig. S11**).

For the survival curves of time from first follow-up to $PCR^+$, we used July 25th as the date of the first follow-up for all individuals and excluded individuals who are already $PCR^+$ at the first follow-up (as in the data).

**Model simulation for a homogeneous population**

The homogeneous population of 1,000 individuals with the same FOI but different age (see **Table S2** and **Fig. S7**) was simulated and analyzed in the same way as the heterogeneous population (with the same parameter values except for the FOI, see **Table S1**). The only difference is that in the homogeneous population all individuals have the same FOI, i.e., instead of the range of biting rates in **Table S1** the biting rate is a fixed constant.

**Heterogeneity in infection risk and the spread of malaria at the beginning of the wet season**

We estimate from the data that carriers have a biting rate of 5.79 bites per year and non-carriers have a biting rate of 1.53 bites per year (**Fig. S13**). Thus, carriers have 3.8 times as many bites per year as non-carriers.

In the following, we aim to quantify how this difference in the biting rate influences the spread of parasites at the beginning of the wet season. Since individuals with negative RDT but positive PCR results ($RDT^-PCR^+$) were not followed-up regularly and we do not have any information on their risk of infection, we assume that their risk of infection is the same as for non-carriers for the following analysis.

In the homogeneous case, where all individuals have the same infection risk, carriers would receive 17.3% of all mosquito bites as 17.3% of the population are carriers (see **Fig. 6**). In the heterogeneous case, where carriers and non-carriers have different biting rates (as estimated from the data), carriers receive the following proportion of mosquito bites:

$$\frac{(\text{biting rate carriers})\times(\text{fraction carriers})}{(\text{biting rate carriers})\times(\text{fraction carriers})+(\text{biting rate non-carriers})\times(\text{fraction non-carriers})},$$

i.e., carriers receive 44.1% of all mosquito bites. Thus, in the heterogeneous case, carriers receive 2.6 times as many bites as in the homogeneous case.

Next, we study how this difference in the fraction of bites that carriers and non-carriers receive influences the spread of parasites in the mosquito-population. To this end, we construct a simple model of the fraction of uninfected mosquitoes decreasing at the rate at which they bite carriers. If there is a baseline biting rate $b$ and the fraction of bites of carriers is $f$, then the fraction of uninfected mosquitoes $m(t)$ decreases in the following way:

$$\frac{d}{dt}\, m(t) = -bf\, m(t), \qquad m(0) = 1.$$

Thus,

$$m(t) = e^{-bf\,t}$$

and the time until a fraction $x$ of the mosquito population is infected is given by

$$t_x = -\frac{\log(1-x)}{bf}.$$

Comparing the time until a fraction $x$ of the mosquito population is infected in the homogeneous and the heterogeneous case, we find that

$$\frac{t_{x,\text{het}}}{t_{x,\text{hom}}} = \frac{f_{\text{hom}}}{f_{\text{het}}} = 0.3919.$$

Thus, in the heterogeneous case it takes only 39.2% of the time to infect a certain proportion of the mosquito population as it would in the homogeneous case or in the homogeneous case it takes 2.6 times as long to infect the same proportion of mosquitoes as in the heterogeneous case.

### Supplementary results

#### Time from enrollment to infection: The effect of age

We found that carriers have a significantly shorter time to infection detected by PCR (see **Fig. 5**h). Here, we also find a significant effect of age with younger children having a longer time to infection (see **Fig. S15**).

Interestingly, the Cox Proportional Hazards model indicates that carriers have a three times higher risk of infection than non-carriers even after accounting for age (**Table S3**). Thus, carriers are more exposed to infectious mosquito bites than non-carriers. As carriers are bitten more, they probably also contribute more to transmission and may be super-spreaders [13].

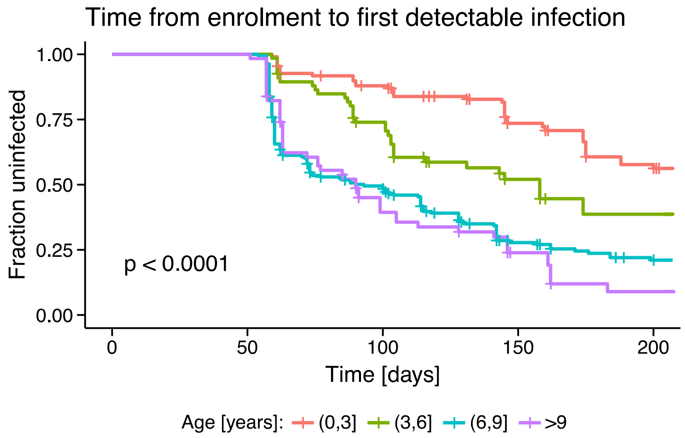


**Fig. S15** Kaplan-Meier curves for the time from enrollment to infection, detected with PCR for carriers, i.e., $RDT^+$ (treated), and non-carriers, i.e., $PCR^-$, and different age groups. The p-values for the comparison of the different survival curves are shown in the lower left corners of the plot.

**Cox PH model for time from enrollment to $PCR^+$**

| Covariate | Coefficient | Exp(Coef) | SE(Coef) | p-value |
|---|---|---|---|---|
| Infection status at enrollment | 1.113 | 3.044 | 0.147 | $3.6\ 10^{-14}$ |
| Age | 0.132 | 1.141 | 0.023 | $8.6\ 10^{-9}$ |

**Table S3** Cox Proportional Hazards model for the time from enrollment to detection of parasites with PCR. The infection status at enrollment is either $RDT^+$ (and thus treated) or $RDT^-$ (untreated). Abbreviations: Exp(Coef): exponential of the coefficient, SE(Coef): Standard Error of the coefficient.

#### Time from first follow-up to infection and clinical malaria

Since there is a gap of approximately two months between enrollment and the first follow-up in the data, we consider the time to infection and clinical malaria from enrollment (see **Fig. 8**) as well as from the first follow-up (see **Fig. S16**). In both cases, carriers have a significantly shorter time to infection but a significantly longer time to clinical malaria than non-carriers.

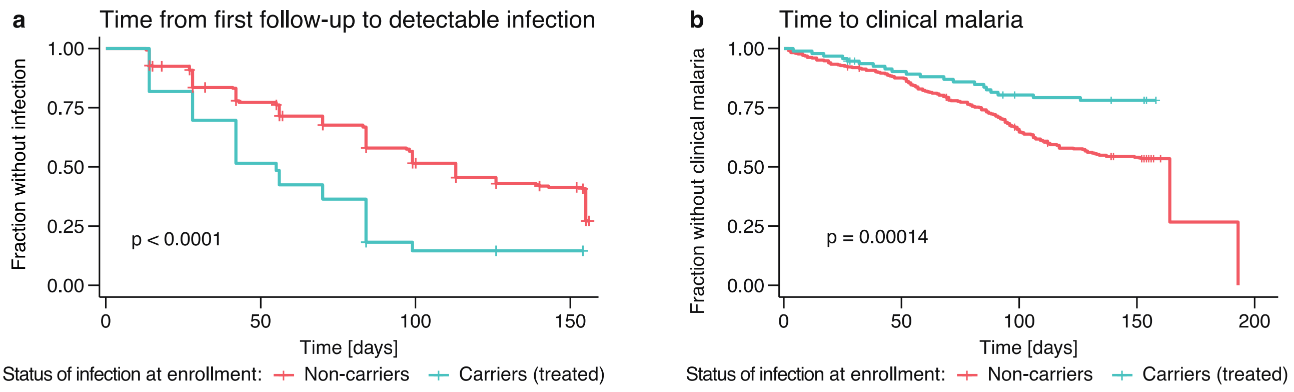


**Fig. S16** Time from the first follow-up visit to infection and to clinical malaria. **a** Time from the first follow-up visit to first detection of parasites with PCR for treated carriers and non-carriers. Carriers have a higher risk of infection compared to non-carriers (log-rank test with p-value < 0.0001). **b** Time from the first follow-up visit to clinical malaria for carriers and non-carriers. Non-carriers have a significantly higher risk of clinical malaria than carriers (log-rank test with p-value 0.00014).

#### Time from first follow-up to infection: The effect of age

We find that the time to the first positive PCR result decreases significantly with age, i.e., the exposure of children increases with age (**Fig. S17** and **Table S4**).

Interestingly, if children and adults aged 4 to 25 years are considered instead of children aged 3 months to 12 years, the age influence is not significantly influencing the risk of infection [14]. The significant influence of age may be due to decreased infection risk for very young children, e.g., because they spend more time under insecticide-treated bed nets. This agrees with the observation that the infection rate increases in the first years of life and then decreases again [15]. If young children (≤ 3 years) are excluded, the effect of age is not significant but the infection status at the end of the dry season still significantly influences the risk of infection (**Fig. S18** and **Table S5**).

It is also interesting to note that in the Cox Proportional Hazards model for the time from first follow-up to PCR$^+$, carriers have only a 1.8 times higher risk of infection than non-carriers (**Table S4**) whereas carriers have a three times higher risk of infection if the time from enrollment to infection is considered. Both models account for age and status of infection at enrollment (carrier or non-carrier), the only difference is that one considers the time from enrollment (late May, i.e., at the end of the dry season) while the other considers time from the first follow-up (approximate two months after enrollment in late July, i.e., after the beginning of the wet season and malaria transmission). This may indicate that the risk of infection and also the contribution of carriers to transmission is higher at the beginning of the wet season than it is during the wet season. However, it may also hint at heterogeneity in the population. If some individuals are more exposed than others, the most exposed are already PCR$^+$ by the time of the first follow-up and thus excluded in the time from first follow-up to PCR$^+$. Thus, the remaining individuals have a lower FOI and we observe a lower risk of becoming PCR$^+$ at a later time point.

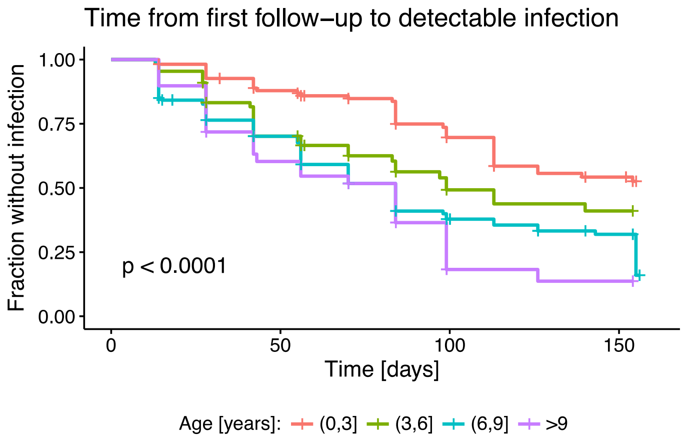


**Fig. S17** Kaplan-Meier curves for the time from first follow-up to positive PCR result for different age groups. The p-value for the comparison of the different survival curves is shown in the lower left corner of the plot. The log-rank test indicates a significant difference in the infection risk for different age groups (p-value < 0.0001).

**Cox PH model for time to detection of parasites with PCR**

| Covariate | Coefficient | Exp(Coef) | SE(Coef) | p-value |
|---|---|---|---|---|
| Infection status at enrollment | 0.607 | 1.834 | 0.218 | 0.005 |
| Age | 0.102 | 1.107 | 0.025 | $3.9\ 10^{-5}$ |

**Table S4** Cox Proportional Hazards model for the time from first follow-up to detection of parasites with PCR. The infection status at enrollment is either RDT$^+$ (and thus treated) or RDT$^-$ (untreated). Abbreviations: Exp(Coef): exponential of the coefficient, SE(Coef): Standard Error of the coefficient.

##### Time from first follow-up to infection: excluding children under 3 years

The increase in exposure of children with age is not significant if children under 3 years of age are excluded (**Fig. S18** and **Table S5**).

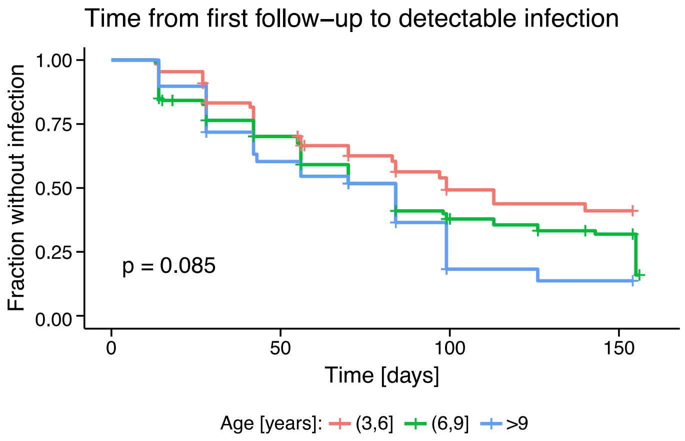


**Fig. S18** Influence of age on the time to first positive PCR result. Kaplan-Meier curves for different age groups and the log-rank test (p-value shown in the lower left corner of the plot) indicate that if children under the age of 3 years are excluded, then there is no significant difference in time to PCR$^+$.

**Cox PH model for time to detection of parasites with PCR (excluding children under 3 years)**

| Covariate | Coefficient | Exp(coef) | SE(coef) | p-value |
|---|---|---|---|---|
| Infection status at enrollment | 0.679 | 1.973 | 0.225 | 0.002 |
| Age | 0.043 | 1.044 | 0.049 | 0.382 |

**Table S5** Cox Proportional Hazards model for the time from first follow-up to detection of parasites with PCR where children under 3 years old are excluded. The infection status at enrollment is either $RDT^+$ (and thus treated) or $RDT^-$ (untreated). Abbreviations: Exp(Coef): exponential of the coefficient, SE(Coef): Standard Error of the coefficient.
